# Cost-effectiveness of screening with transcriptional signatures for incipient TB among U.S. migrants

**DOI:** 10.1101/2024.10.09.24315062

**Authors:** Yuli Lily Hsieh, C Robert Horsburgh, Ted Cohen, Jeffrey W Miller, Joshua A Salomon, Nicolas A Menzies

## Abstract

**Introduction:** Host-response-based transcriptional signatures (HrTS) have been developed to identify “incipient tuberculosis (TB)”. No study has reported the cost-effectiveness of HrTS for post-arrival migrant screening programs in low-incidence countries.

**Objectives:** To assess the potential health impact and cost-effectiveness of HrTS for post-arrival TB infection screening among new migrants in the United States.

**Methods:** We used a discrete-event simulation model to compare four strategies: (1) no screening for TB infection or incipient TB; (2) ‘IGRA-only’, screen all with interferon gamma release assay (IGRA), provide TB preventive treatment for IGRA-positives; (3) ‘IGRA-HrTS’, screen all with IGRA followed by HrTS for IGRA-positives, provide incipient TB treatment for individuals testing positive with both tests; and (4) ‘HrTS-only’, screen all with HrTS, provide incipient TB treatment for HrTS-positives. We assessed outcomes over the lifetime of migrants entering the U.S. in 2019, assuming HrTS met the WHO Target Product Profile (TPP) optimal criteria. We conducted sensitivity analyses to evaluate the robustness of results.

**Results:** The IGRA-only strategy dominated the HrTS-based strategies under both healthcare sector and societal perspectives, with an incremental cost-effectiveness ratio of $78,943 and $89,431 per quality-adjusted life-years (QALY) gained, respectively. This conclusion was robust to varying costs ($15–300) and characteristics of HrTS, and the willingness-to-pay threshold ($30,000–150,000/ QALY gained), but sensitive to the rate of decline in TB progression risk after U.S. entry.

**Conclusions:** Our findings suggest that HrTS meeting the WHO TPP is unlikely to be a cost-effective component of post-arrival screening for migrants entering the U.S.

## Introduction

Without preventive treatment, approximately 5-10% of healthy individuals infected with *Mycobacterium tuberculosis (Mtb)* will progress to tuberculosis (TB) disease during their lifetime.^1,2^ Current WHO guidelines recommend targeting TB preventive treatment to infected persons at highest risk of disease progression. However, current tests for *Mtb* infection, including the interferon gamma release assay (IGRA) and tuberculin skin test (TST), have low positive predictive values for the proximal onset of TB disease, with only 1-6% of individuals identified with these tests developing TB within two years.^3–6^ The lack of tools for predicting TB disease limits the ability of TB programs to target preventive treatment to those at highest risk.

In recent years, host-response-based transcriptional signatures (henceforth, HrTS) have been investigated for their potential to identify “incipient TB” based on the association between changes in host transcriptome and risk of disease progression.^7,8^ Incipient TB has been defined as “*Mtb* infection that is likely to progress to TB disease in the absence of further intervention but has not yet induced clinical symptoms, radiographic abnormalities, or microbiologic evidence consistent with TB disease”.^9^ If HrTS can distinguish incipient TB from non-progressing *Mtb* infection, these tests could facilitate more targeted delivery of interventions to prevent disease progression. WHO has established a Target Product Profile (TPP) for this class of tests, recommending sensitivity and specificity of at least 75% (minimum criteria), and ideally 90% (optimal criteria), for distinguishing individuals who would progress to TB disease within two years.^7^

One potential use case of HrTS is for targeting TB preventive treatment among migrants from countries with high TB incidence moving to lower TB incidence settings. In many low-incidence countries, migrants have elevated TB incidence rates as a result of infection acquired before or during migration.^10^ For example, in the United States, 71% of reported TB cases between 2017 and 2021 occurred in non-US-born persons,^11^ and most were attributable to infection acquired before entry.^12^ Screening and treatment of *Mtb* infection in high-risk migrant populations has been identified as a key component in global tuberculosis control effort.^13^ In this setting, HrTS may be useful as a rule-out test to reduce the need to treat all individuals with a positive IGRA result, the majority of whom are at very low risk of proximal progression. It could also be used as a stand-alone rule-in test for incipient TB. To the best of our knowledge, no study has evaluated the cost-effectiveness of such a test as a post-arrival screening tool for *Mtb* infection among migrants to low-TB burden countries.

In this study, we assessed the potential health impact and cost-effectiveness of four post-arrival screening strategies for *Mtb* infection among new U.S. migrants, using a potential HrTS that meets the WHO TPP optimal criteria. To compare these strategies, we used a decision analytic framework with a discrete-event simulation (DES) model. We parameterized this model with data for the 2019 immigrant cohort to estimate lifetime TB-related health outcomes and health service utilization under each screening scenario.

## Methods

### Study Population

Our simulated study cohort (n = 2,042,225) included migrants whose annual TB risk could be estimated from a published TB risk model as a function of migrants’ country-of-origin, entry year, age at entry, and number of years since entry to the U.S.^14^ Our study cohort represented 97 countries-of-origin, accounting for 70.6% of the 2019 entry cohort and the majority of TB cases among this population. The population sizes by age and country-of-origin were estimated from 2019 American Community Survey data. We divided the study population into four risk categories based on WHO-estimated country-of-origin TB incidence rates in 2019 (I, 0-9.9; II, 10-99.9; III, 100-299.9; and IV, ≥300 per 100,000). We conducted our analyses for the entire study cohort and by risk category (Table 1). Further details are given in Appendix 1 in the online supplement.

**Table 1.** Characteristics of study cohort upon entry to the US in 2019.

| Risk category<sup>1</sup> | Number of countries/<br>regions | Countries/ regions<sup>2</sup> | Modelled population<br>size, n<br>(% of total population) | Mean entry<br>age |
| --- | --- | --- | --- | --- |
| Whole cohort | 97 |  | 2,042,225 (100 %) | 29.0 |
| I | 5 | IDN, LBR, MMR, **PHL**, ZAF | 100,778 (4.9 %) | 34.0 |
| II | 19 | AFG, BGD, BOL, CMR, ETH, GHA,<br>HTI, **IND**, KEN, KHM, LAO, NGA,<br>NPL, PAK, PER, SLE, SOM, THA,<br>**VNM** | 340,454 (16.7 %) | 28.9 |
| III | 50 | ALB, ARG, ARM, BGR, BIH, BLR,<br>BLZ, BRA, CHL, **CHN**, COL, CPV,<br>CRI, DOM, ECU, EGY, ESP, FJI,<br>GTM, GUY, HKG, HND, IRN, IRQ,<br>JPN, KOR, LBN, LKA, LTU, MAR,<br>MDA, **MEX**, MYS, NIC, PAN, POL,<br>PRT, ROU, RUS, SDN, SLV, SYR,<br>TTO, TUR, TWN, UKR, URY, UZB,<br>VEN, YEM | 1,409,531 (69.0 %) | 27.9 |
| IV | 23 | AUS, AUT, BEL, BRB, CAN, CHE,<br>CUB, CZE, DEU, FRA, GBR, GRC,<br>GRD, HRV, HUN, IRL, ISR, ITA, JAM,<br>JOR, NLD, SAU, SWE | 191,462 (9.4 %) | 34.7 |

<sup>1</sup> Epidemiological categorization of migrant populations based on TB incidence per 100k in 2019 for their country-of-origin: risk category I ( $\geq 300$ ); risk category II (100-300), risk category III (10-100), risk category IV (0-10).
<sup>2</sup> Countries highlighted in red are the top five country-of-origins that contribute to the total number of TB cases among migrants.

### Intervention Strategies

We compared four post-arrival screening strategies: (I) no screening for TB infection or incipient TB (‘no screening’); (II) screen all with IGRA, provide TB preventive treatment for individuals testing positive (‘IGRA-only’); (III) screen all with IGRA followed by HrTS for IGRA-positive persons, provide incipient TB treatment for individuals testing positive with both tests (‘IGRA-HrTS’); and (IV) screen all with HrTS, provide incipient TB treatment for individuals testing positive (‘HrTS-only’). Screening was assumed to occur one month after U.S. arrival. Strategies II - IV also assumed screening and treatment for TB disease prior to provision of TB preventive treatment or incipient TB treatment. We assumed that, if diagnosed, TB disease would be treated with a standard first line treatment, *Mtb* infection would be treated with once-weekly isoniazid-rifapentine for 12 weeks, and incipient TB would be treated with one month of daily isoniazid-rifampicin followed by three months of isoniazid-rifampicin thrice weekly.^15^ Table 2 summarizes clinical decision rules, and Appendix 2 in the online supplement provides a flowchart for each strategy.

**Table 2.** Testing and treatment decision rules.

| Tests performed | Test results<sup>1</sup> |  |  | Diagnosis, conditional on test results | Treatment prescribed, conditional on diagnosis<sup>2</sup> |
| --- | --- | --- | --- | --- | --- |
|  | IGRA | HrTS | TB diagnosis |  |  |
| Strategy II<br>(IGRA → TB diagnosis) | pos | -- | pos | TB disease | Treatment for TB disease |
|  | pos | -- | neg | *Mtb* infection | Treatment for *Mtb* infection |
|  | neg | -- | -- | No *Mtb* infection | No treatment |
| Strategy III<br>(IGRA → HrTS → TB diagnosis) | pos | pos | pos | TB disease | Treatment for TB disease |
|  | pos | pos | neg | Incipient TB | Treatment for incipient TB |
|  | pos | neg | -- | Non-progression *Mtb* infection | No treatment |
|  | neg | -- | -- | No *Mtb* infection | No treatment |
| Strategy IV<br>(HrTS → TB diagnosis) | -- | pos | pos | TB disease | Treatment for TB disease |
|  | -- | pos | neg | Incipient TB | Treatment for incipient TB |
|  | -- | neg | -- | No incipient TB | No treatment |
| In all strategies, in the case of TB diagnosis that occurred outside of the post-arrival screening | -- | -- | pos | TB disease | Treatment for TB disease |

<sup>1</sup> pos, positive; neg, negative; TB diagnosis is assumed to be perfect.
<sup>2</sup> In this study, we assumed individuals were given 3HP for treatment for *Mtb* infection and one month of HR daily followed by three months of HR three times a week for treatment for incipient TB, a regimen that has been used for sputum negative TB cases.<sup>1</sup>

For IGRA, we defined sensitivity and specificity with respect to the *Mtb* infection status at the time of testing. We assumed IGRA would be positive in 98.5% of individuals with *Mtb* infection and negative in 78.9% of individuals without *Mtb* infection.^16^ For HrTS, we defined sensitivity and specificity of HrTS with respect to prevalent and future TB disease. For specificity, we assumed that HrTS would be negative in 90% of individuals without infection or with infection that would not lead to disease before death. We assumed HrTS would have a sensitivity of 90% for prevalent TB as well as TB cases occurring within two years of testing, and a sensitivity of 10% for cases occurring subsequently. This assumption implies the test cannot distinguish individuals who will from those who will not develop TB disease beyond the two-year predictive period, consistent with the reported performance of current signatures.^17^ Further, we assumed independence in IGRA and HrTS test results, conditional on an individual’s TB-related health status at the time of testing ((1) no *Mtb* infection, (2) *Mtb* infection that will not progress to TB disease during lifetime, (3) *Mtb* infection that will progress within two years (incipient TB), (4) *Mtb* infection that will progress after two years, (5) prevalent TB disease).

### Discrete Event Simulation (DES) Model

Each migrant in the study population was represented as an individual in the DES model, characterized by country-of-origin, entry age, and initial health state upon U.S. arrival. The three possible initial health states were: (1) no *Mtb* infection, (2) *Mtb* infection without TB disease, (3) TB disease. Initial health states were assigned based on estimated prevalence of *Mtb* infection and estimated incidence of TB disease, by age and country-of-origin. To parameterize progression from *Mtb* infection to TB disease, we estimated individual TB risk over time by applying the demographic data of the 2019 entry cohort to the fitted TB risk model reported in Hill *et al*.^14^ This empirical study estimated TB incidence rates among U.S. migrants after entry to the country, as a function of entry year, entry age, country-of-origin, and time since entry (Appendix 1 in the online supplement).

To estimate *Mtb* infection prevalence of by country-of-origin and age group, we fit a functional relationship between *Mtb* infection prevalence and TB incidence rate using data reported by the U.S. TB Epidemiological Studies Consortium,^18^ and calibrated this to match the overall prevalence of *Mtb* infection in the non-US-born population.^19^ See Appendix 3 in the online supplement for details. The fraction of migrants entering the US with TB disease was based on the number of TB cases diagnosed in the 6 months following U.S. entry, estimated using the published TB risk model.^14^

At the beginning of the simulation, each individual was assigned a time-to-TB value, drawn randomly from empirical survival functions of TB incidence, described in Appendix 4 in the online supplement. Each individual was also assigned a time-to-death value, drawn from survival functions derived from the 2017 U.S. Life Tables for non-US-born individuals.^20^ Whether a simulated individual would develop TB disease in their lifetime was determined by the Gillespie algorithm:^21^ in the absence of intervention, those with a time-to-TB shorter than the time-to-death were expected to develop TB during their lifetime. Event data from U.S. entry to death were simulated and recorded for all modelled individuals. The model was developed in R.^22^ Appendix 5 in the online supplement reports parameter values.

### Positive predictive value and negative predictive value

For each screening strategy, we calculated the positive predictive value (PPV) and negative predictive value (NPV) of ever having TB in their lifetime, including prevalent TB disease at the time of testing. Additionally, we estimated the PPV and NPV of having TB disease within two years of testing, including prevalent TB disease.

### Health outcomes and economic evaluation

We estimated the number of TB cases averted, quality adjusted life years (QALYs) gained, number of each test administered, and number of each treatment prescribed under each strategy. In the base case scenario, IGRA and HrTS were $62 and $30 in 2021 U.S. dollars, respectively. Other healthcare related and non-healthcare related costs are in Appendix 5 in the online supplement. We conducted a cost-effectiveness analysis from both societal and healthcare sector perspectives, with costs and QALYs discounted at 3% annually.^23^ We also calculated the net monetary benefit (total monetized health benefits minus total societal costs) for each strategy with health gains valued at US$150,000 per QALY,^24^ and other values considered in sensitivity analyses. Appendix 7 in the online supplement provides an impact inventory and additional costing methods.

### Sensitivity Analysis

We conducted probabilistic sensitivity analysis (PSA) to determine the robustness of our results to collective uncertainty in all parameter inputs, generating 1000 simulated values for all cohort outcomes.^25^ The distributions of parameters included in the PSA are in Table E3 in the online supplement. We conducted an additional two-way sensitivity analysis over the cost of HrTS (US$15 – $300) and willingness-to-pay threshold ($30,000 – $150,000 per QALY gained). We also performed a multi-way sensitivity analysis across plausible ranges of sensitivity (75 – 100%), specificity (75 – 100%), and cost of HrTS ($15 – $300), along with the timeframe over which the sensitivity value holds (2 – 10 years) and the willingness-to-pay threshold. The multi-way sensitivity analysis helps inform whether and under what circumstances the HrTS strategies would be cost-effective. Finally, we estimated results for an alternative scenario that assumed a more rapid decline in the rate of progression to TB disease with increasing years since U.S. entry. In this scenario, the annual number of TB cases remained the same as in the main analysis, but the proportion of cases attributable to a pre-existing infection was assumed to decline by 3.4% with each additional year since entry. The remainder were assumed to result from TB (re)infection after U.S. entry, and could not be averted by the modelled interventions. See Appendix 10 for details.

Point estimates for all reported outcomes were calculated as the mean of the 1000 values generated by the PSA, with uncertainties expressed in 95% credible intervals, unless otherwise specified. The incremental cost-effectiveness ratio (ICER) was calculated as the ratio of the mean of incremental cost to the mean of the incremental QALY gained.

## Results

### Expected number of TB cases identified by years since entry

The median age at entry of the study cohort was 29 years old, and the estimated LTBI prevalence was 12.4% (95% credible interval (CI), 8.5 to 43.5). Under the no-screening strategy, we projected 5601 (3667 to 8177) members of the study cohort would develop TB over their lifetime (2.7 per 1000, 2.4 to 3.1). For these individuals, the expected median time to TB was estimated to be 11.3 (IQR 3.1, 26.3) years, with 20.2% (18.6 to 21.7) of cases occurring within two years of U.S. arrival. The CDC Online Tuberculosis Information System Data reported 840 TB cases among non-US-born population within the first year they entered the U.S. in 2019, and our risk model estimated 770 (95% CI 629 to 894) cases.^18^

Figure 1 shows the projected number of TB cases by year under each strategy. In the no-screening and the IGRA-only strategies, the trend decreases monotonically. In contrast, the IGRA-HrTS and the HrTS-only strategies show a slight rebound after year one followed by a monotonic decrease thereafter, as HrTS identified most of the incipient TB at the time of screening. For the IGRA-HrTS and HrTS-only strategies, TB incidence reductions are concentrated in the years following screening, while the TB incidence reduction accumulates over a longer period, proportional to overall incidence trends, in the IGRA-only strategy.

**Figure 1.**
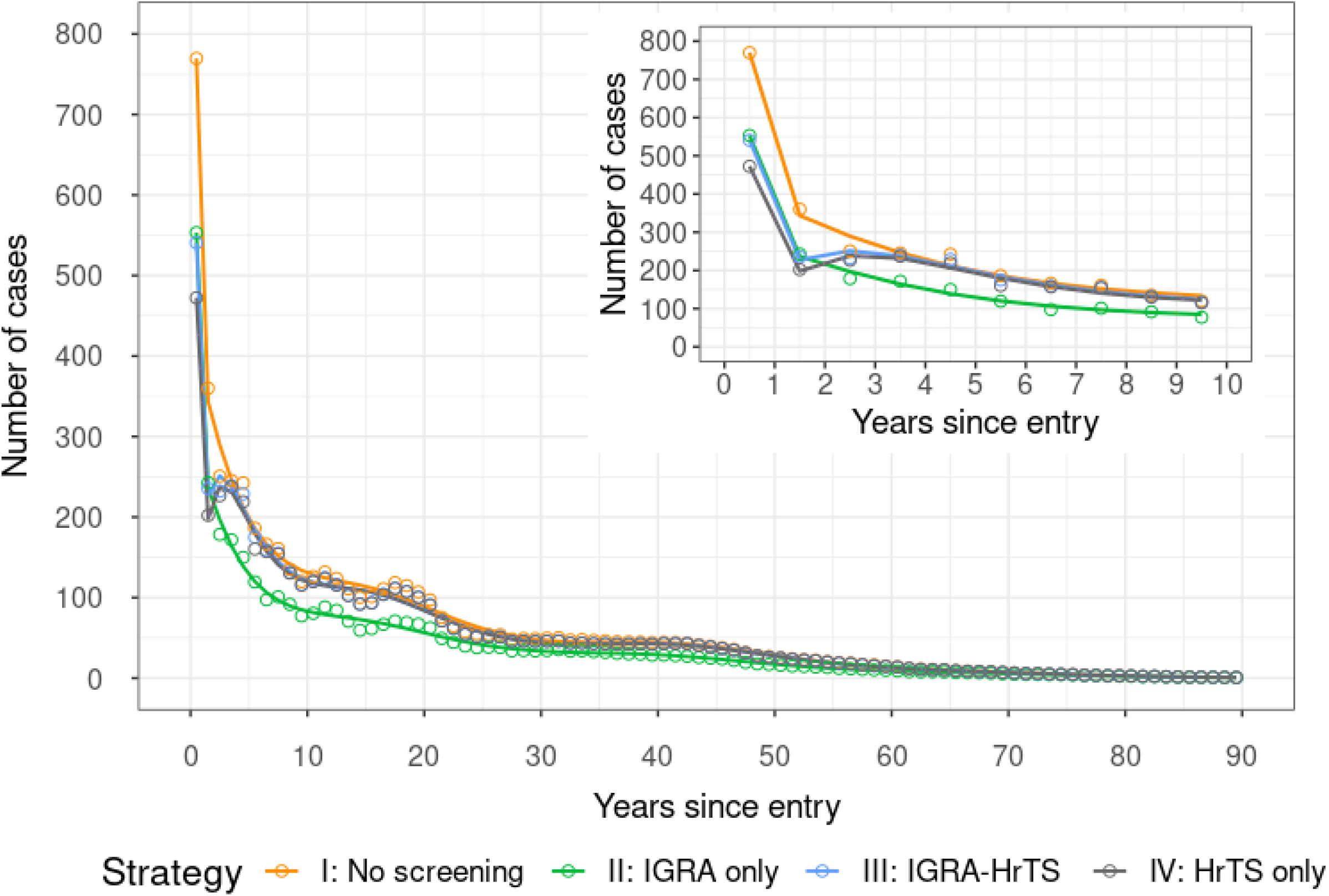
Number of TB cases, by years since entry. The circles represent the expected number of TB cases in each year, and the lines are fitted smooth lines to show the trend.

Compared to the no-screening strategy, the IGRA-only strategy was estimated to result in the greatest reduction (33.7%, 25.7 to 42.8) in TB cases over the lifetime of the entire study cohort, followed by the HrTS-only strategy (13.2%, 11.3 to 16.0) and the IGRA-HrTS strategy (10.4%, 9.0 to 12.2) (Table 3). This pattern was consistent across risk categories.

**Table 3.**
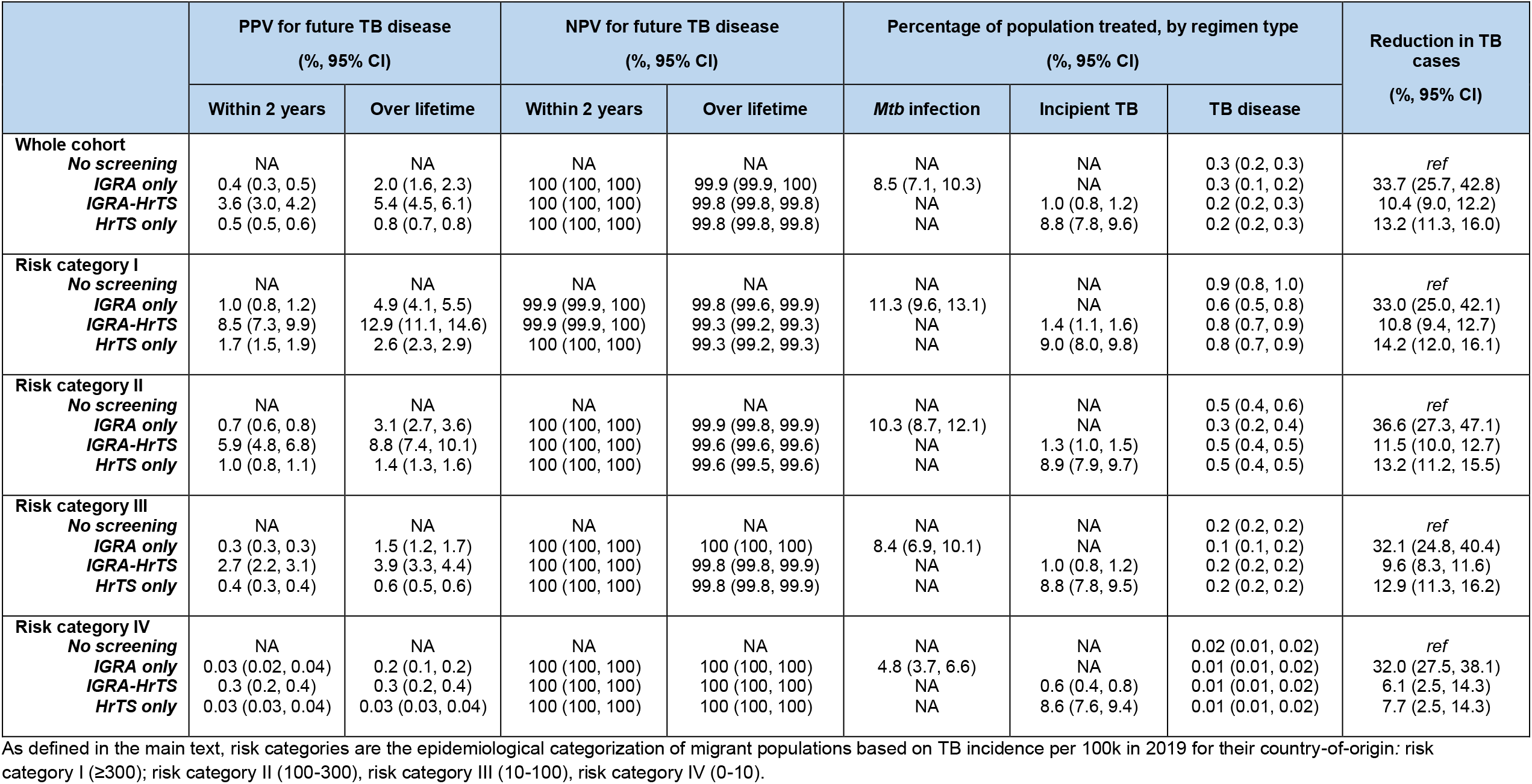
Testing and treatment outcomes.

### Utilization of screening and treatment resources

In the IGRA-only strategy, 11.1% (95% CI 9.2 to 13.4) of the study cohort tested positive with IGRA, 1.1% (95% CI 0.9 to 1.4) tested positive with both IGRA and HrTS in the IGRA-HrTS strategy, and 10.0% (95% CI 9.99 to 10.02) tested positive with HrTS in the HrTS-only strategy (Online Supplement Table E4). The proportions of the cohort receiving an intervention to prevent disease progression were more than eight times higher in the IGRA-only strategy (8.5%, 7.1 to 10.3) and the HrTS-only strategy (8.8%, 7.8 to 9.6), compared to the IGRA-HrTS strategy (1.0 %, 0.8 to 1.2) (Table 3).

The expected proportions treated for TB disease, identified through active or passive screening, were highest in the no-screening strategy, followed by the IGRA-HrTS strategy, and the IGRA-only strategy (Table 3).

### PPV and NPV of screening algorithms

Overall, the PPV for TB disease within two years (3.6%, 3.0 to 4.2) and over the lifetime (5.4%, 4.5 to 6.1) were highest in the IGRA-HrTS strategy. The PPV for TB disease within two years was the lowest in the IGRA-only strategy (0.4%, 0.3 to 0.5) and the PPV for TB disease over lifetime was the lowest in the HrTS-only strategy (0.8%, 0.7 to 0.8). Comparing across risk groups, PPV values were higher among migrant populations from countries with higher TB risks (Table 3). The NPV for future TB disease within two years and over lifetime were greater in lower risk categories, but were high overall for all strategies (Table 3).

### Health benefits, costs, and cost-effectiveness

Relative to the no-screening strategy, the IGRA-only strategy produced the greatest per-person gain in QALYs whereas the IGRA-HrTS strategy offered the least. From the healthcare sector perspective, the HrTS-only strategy incurred the greatest additional costs while the IGRA-only strategy incurred the least. From the societal perspective, the HrTS-only strategy was the costliest, while the IGRA-HrTS strategy was the least costly. Compared to the no-screening strategy, the IGRA-only strategy resulted in an ICER of US$78,943 per QALY gained in the healthcare sector perspective and an ICER of $89,431 in the societal perspective, dominating the IGRA-HrTS and HrTS-only strategies (Table 4). The subgroup cost-effectiveness results are in Appendix 8 in the online supplement.

**Table 4.** Reference case cost-effectiveness results (time horizon: lifetime; costs and health effects incremental to the ‘no screening’ strategy, discounted at 3% annually)

| Strategy | TB related<br>HC costs | Other HC<br>expenditures | TB related<br>non-HC<br>costs | Other non-HC<br>expenditures | Productivity<br>gain<sup>1</sup> | Total Costs<sup>2</sup><br>\$ | Total QALY gain | Inc. Cost<sup>2</sup> | Inc.<br>Effectiveness<sup>3</sup><br>(QALYs) | ICER | Inc.<br>NMB<sup>4</sup> |
| --- | --- | --- | --- | --- | --- | --- | --- | --- | --- | --- | --- |
| **Healthcare Sector Perspective** |  |  |  |  |  |  |  |  |  |  |  |
| **IGRA only** | 92.4<br>(67.4, 121.7) | 4.6<br>(2.4, 6.7) | -- | -- | -- | 97.1<br>(72.4, 126.4) | 0.00123<br>(0.00083, 0.00168) | 97.1 | 0.00123 | 78,943 | 87.4 |
| **IGRA-HrTS** | 71.1<br>(55.4, 89.0) | 1.1<br>(0.6, 1.7) | -- | -- | -- | 72.2<br>(56.4, 90.2) | 0.00082<br>(0.00048, 0.00120) | NA | NA | Extended<br>dominated | 50.8 |
| **HrTS only** | 75.4<br>(55.5, 101.7) | 1.3<br>(0.7, 2.0) | -- | -- | -- | 76.7<br>(56.8, 103.0) | 0.00077<br>(0.00038, 0.00118) | NA | NA | Dominated | 38.8 |
| **Societal Perspective** |  |  |  |  |  |  |  |  |  |  |  |
| **IGRA only** | 92.4<br>(67.4, 121.7) | 4.6<br>(2.4, 6.7) | 6.8<br>(4.4, 9.6) | 16.2<br>(8.1, 23.6) | 10.1<br>(4.5, 14.8) | 110.0<br>(85.3, 140.1) | 0.00123<br>(0.00083, 0.00168) | 110.0 | 0.00123 | 89,431 | 74.5 |
| **IGRA-HrTS** | 71.1<br>(55.4, 89.0) | 1.1<br>(0.6, 1.7) | 1.1<br>(0.4, 1.7) | 4.9<br>(2.6, 7.2) | 4.7<br>(1.4, 8.3) | 73.4<br>(57.6, 91.8) | 0.00082<br>(0.00048, 0.00120) | NA | NA | Extended<br>Dominated | 49.6 |
| **HrTS only** | 75.4<br>(55.5, 101.7) | 1.3<br>(0.7, 2.0) | 12.5<br>(9.9, 15.3) | 5.6<br>(2.8, 8.4) | 5.2<br>(1.7, 9.0) | 89.6<br>(69.2, 115.9) | 0.00077<br>(0.00038, 0.00118) | NA | NA | Dominated | 25.9 |

Notes: HC, healthcare; QALY, quality-adjusted life years; ICER, incremental cost-effectiveness ratio; NMB, net monetary benefit
1 All costs, expenditures, productivity gain, and QALY gain were estimated relative to the “No Screening” strategy.
2 Productivity gain was attributable to mortality aversion.
3 For analysis in the healthcare sector perspective, Total Costs = (TB related healthcare costs + Other healthcare expenditures); for analysis in the societal perspective, Total Costs = (TB related healthcare costs + Other healthcare expenditures + TB related non-healthcare costs + Other non-healthcare expenditures – Productivity gain)
4 The Inc. cost and Inc. effectiveness in columns 9-10 were estimated relative to the next best strategy.
5 The NMB were based on \$150,000/QALY; Inc. NMB = (Inc. effectiveness\* \$150,000/QALY) – Inc. costs

### Sensitivity analysis

Two-way sensitivity analyses show that our cost-effectiveness analysis conclusion is robust to the cost of HrTS as well as the willingness to pay threshold (Online Supplement Figures E2-1, E2-2), even when the cost of HrTS is as low as $15. Our four-way sensitivity analyses show that for the highest risk group, when the cost of HrTS is $15, the HrTS-only strategy may be cost-effective from the societal perspective when HrTS has a >75% sensitivity for TB cases occurring seven years after testing, or nine years when analyzed from the healthcare sector perspective (Online Supplement Figures E3-1-1, E3-1-2).

In an alternative scenario that assumed a more rapid decline in the rate of progression to TB disease following U.S. entry, the IGRA-only strategy remained the cost-effective strategy from both healthcare sector and societal perspectives, at a $150,000 willingness-to-pay threshold and a $30 HrTS. However, at a willingness-to-pay threshold to $100,000 the IGRA-HrTS strategy would be cost-effective from the healthcare sector perspective (Online Supplement Table E11).

## Discussion

To our knowledge, this is the first analysis to formally evaluate the cost-effectiveness of incorporating host transcriptomic signatures in post-arrival screening algorithms among migrants in a low-incidence country. In our main analysis, we found that, compared to the current recommendation of using IGRA to screen for and treat *Mtb* infection, it would not be cost-effective to use HrTS either as a rule-out test among IGRA positives or as a stand-alone rule-in test for incipient TB among newly arrived migrants in the United States, even if these tests meet the WHO TPP optimal criteria. In our study, HrTS was assumed to have a 90% sensitivity of TB cases occurring in the first two years after U.S. arrival, which represented 20% of TB cases occurring over the lifetime of our study cohort. HrTS only had a 10% sensitivity for the remaining cases. In contrast, while IGRA had a lower sensitivity for incipient cases, these tests retained a 78% sensitivity for cases occurring after this initial two-year period, a key reason why the IGRA-only strategy dominated the other screening strategies.

If HrTS could predict more cases with delayed onset, these new screening tools may be cost-effective. As we report in sensitivity analyses, the HrTS-only strategy would be preferred if HrTS retains at least a 75% sensitivity for TB cases occurring over at least the subsequent seven years. However, it is unlikely that HrTS signatures with high sensitivity over an interval longer than two years are imminent.^17^ In addition, in an alternative scenario that assumed more rapid declines in the rate of progression to TB disease following U.S. entry, the health benefits estimated for the IGRA-only strategy were proportionally lower, and HrTS was found to be potentially cost-effective. In this analysis, the IGRA-HrTS strategy became the preferred strategy from the healthcare sector perspective for a willingness-to-pay threshold for $100,000 per QALY, indicating that the cost-effectiveness of HrTS depends on the long-term health benefits achieved with the current IGRA-only strategy. Additional studies estimating the dynamics of TB progression risk many years after migration would be valuable.

We have evaluated only one specific use case of HrTS among recent migrants to low incidence settings, and HrTS may have greater value in settings with higher TB incidence. A modelling study by Sumner *et al*. showed that targeted TB preventive therapy guided by a blood transcriptomic biomarker (RISK11) may be more effective in averting TB cases than a one-off universal treatment amongst people living with HIV but would require repeat testing.^26^ The modelled cohort they evaluated had a TB incidence rate much higher than in our study. They also reported that for the signature to be cost-effective, the cost of the test would need to be one-tenth of the preventive therapy regimen, assuming annual screening. HrTS could also be used to screen for active TB disease in migrant populations. However, a cost-effectiveness analysis of HrTS for TB screening in adults with suspected TB disease in the United Kingdom showed that it would not be cost-effective compared to the status quo.^27^

We found that the IGRA-only strategy had the lowest PPV for incipient TB disease while the HrTS-only strategy had the lowest PPV for TB disease over the remaining lifetime. This reflects the assumption that after the initial two-year window, the HrTS has a 10% false positive rate, substantially higher than the 1.5% with IGRA. This result highlights that both the PPV and NPV are important metrics for quantifying the overall value provided by these predictive tests. Compared to the PPV of IGRA reported in the UK PREDICT TB study (3.0%), our study found a lower PPV of IGRA (0.4% for TB within two years). However, in our study population, the TB incidence rate was 38 per 100,000 person-years in year 1 and 18 per 100,000 in year 2; in the PREDICT study, incidence was 932 per 100,000 in year 1 and 115 per 100,000 in year 2. Given that the PPV will be sensitive to the incidence rate in the population of interest, this difference in TB risk could explain the difference in PPV.

Our study has several strengths. First, we used an individual based model with granular, time-dependent TB risk estimates that are a function of each migrant’s age, entry-year, country-of-origin, and years since U.S. arrival. This allowed us to estimate lifetime outcomes of the study cohort under different scenarios and evaluate the impact of HrTS in a real-world setting that has direct clinical and policy implications. The fact that the analysis captured lifetime outcomes is particularly important because HrTS is a predictive test for future TB disease; restricting the analytic timeframe could over-estimate the comparative effect of it relative to other screening tools.

Second, our analytic framework can be used to evaluate the performance of other predictive tests with sensitivity dependent on individual patient’s time to disease in a setting where the prevalence of a disease is also time-varying. Our approach showed that an individual-based discrete event simulation model has the flexibility to incorporate these two time-dependent elements. Third, our functional definitions of a ‘positive test’, a ‘negative test’, and a ‘TB case’ in PPV and NPV calculations provided a clear performance measure for a predictive test for future TB disease. This can be useful in evaluation of novel screening and diagnostic tools as the field has an increasing interest in tools that can identify individuals who are progressing from *Mtb* infection to TB disease.

One limitation in our study is that we assumed conditional independence of IGRA and HrTS test results. If this assumption does not hold, we may over-estimate the sensitivity of the IGRA-HrTS strategy. We estimated country-specific LTBI prevalence among migrants based on a previously published study that focused on high-risk populations, which might not be representative of the general migrant population. To address this limitation, we calibrated the estimates so that the LTBI prevalence of the entire migrant population in our study matched the national non-US born LTBI prevalence estimate reported in NHANES. Other key assumptions we made in our study included the treatment uptake and completion rates and regimen efficacy for incipient TB. Screening and treatment of incipient TB is not routine clinical practice, and there is little empirical evidence to inform these estimates.

Many signature tests have been proposed in recent years, with the goal of identifying individuals with a high proximal risk of developing TB disease.^3,17^ Currently, none has met the WHO TPP optimal criteria. Our study suggests that even if the WHO TPP criteria were met, HrTS is unlikely to be cost-effective compared to the conventional IGRA test as a post-arrival screening tool among high-risk migrants in our setting.

## Data Availability

All data produced in the present study are available upon reasonable request to the authors.

## Author contributions

YLH, NAM, CRH, TC, and JAS conceived the study. YLH developed the study approach. NAM, CRH, TC, and JAS helped refine the study objectives and study approach. NAM helped gain access to the TB risk model data. YLH and NAM had access to all the data in the study. YLH performed the statistical and modelling analyses. All authors contributed to the interpretation and presentation of study results. YLH wrote the first draft of the manuscript and all authors reviewed, edited, and approved the final version of the manuscript. All authors had final responsibility for the decision to submit for publication.

## Acknowledgements

We thank Dr. Andrew Hill for sharing the detailed results of the TB risk model reported in the study “Hill AN, *et al*. High-resolution estimates of tuberculosis incidence among non-U.S.-born persons residing in the United States, 2000–2016. *Epidemics* 2020; **33**: 100419”, which was funded by the U.S. Centers for Disease Control and Prevention, National Center for HIV/AIDS, Viral Hepatitis, STD, and TB Prevention Epidemiologic and Economic Modeling Agreement (#5NU38PS004644). We also thank Carla Winston, Michelle Van Handel, Julie Self, Suzanne Marks, and Garrett Asay from the U.S. CDC for their valuable input to our manuscript. The findings and conclusions in this paper are those of the authors and do not represent the view of the funding sources or the authors’ affiliated institutions.

## Data Sharing

Analytic code and model inputs are available through the following link: << Link to GitHub repository removed for peer review. It will be made public when the manuscript is accepted and finalized >>. The detailed results of the TB risk model are available at << Dataverse link will be added when the manuscript is accepted and finalized >>.

