## Supplementary material for "Cost-effectiveness of screening with transcriptional signatures for incipient TB among U.S. migrants": Online Supplement

#### Appendix 1

##### Construction and characteristics of the study cohort

Among all the migrants who entered the U.S. in 2019, we included migrants whose annual TB risks could be estimated from the fitted TB risk model reported in a prior study of TB incidence rates in the non-US-born residents of the United States (Hill *et al.*).[1] In this study, the authors constructed generalized additive regression models to estimate TB incidence rates as a function of birth country, entry year, age at entry, and number of years since entry to the United States. They trained the model with individual-level data from the National Tuberculosis Surveillance System (NTSS) on TB cases among non-US-born individual between 2000-2016, and population data from the American Community Survey (ACS) and 2000 U.S. Census. This original study was funded by the U.S. Centers for Disease Control and Prevention, National Center for HIV/AIDS, Viral Hepatitis, STD, and TB Prevention Epidemiologic and Economic Modeling Agreement (#5NU38PS004644). The regression output was made available to the authors of this current project as an `.rdata` object. The R object is available at <<Dataverse link to be added upon acceptance once analyses finalized>>.

The Hill *et al*. model can be used to estimate TB risk for migrants from top 100 countries of birth by U.S. population size between 2000 and 2016. However, among the 100 countries, Yugoslavia (YUG) and the Soviet Union (SUN) no longer existed in 2019 and hence excluded from our analysis. Further, we used smoothed estimates from the 2019 ACS data to inform the population size by country-of-origin and by age (ranged 0-91 years; those >= 92 years old were recoded as 91-year-olds) to adjust for undercounting due to sampling timing of the year. However, we did not have estimates of the population size for migrants for Denmark (DNK) by age, so Denmark was excluded as well. Hence, our study cohort consisted of migrants from the top 97 country-of-origins by U.S. population.

**Table E1. Characteristics of study cohort upon entry to the US in 2019 by country-of-origin**

| Countries/regions | Modelled population size (n) | Mean entry age (years) | Estimated LTBI prevalence % (95% CI) |
|---|---|---|---|
| SOM | 388 | 32.41 | 22.49 (9.08, 44.30) |
| LBR | 548 | 32.25 | 21.28 (9.18, 44.05) |
| ETH | 4651 | 25.95 | 20.94 (9.00, 44.41) |
| MMR | 2580 | 26.47 | 20.68 (9.13, 44.11) |
| KEN | 3781 | 24.98 | 20.27 (9.07, 44.29) |
| SLE | 216 | 23.21 | 20.07 (9.24, 43.68) |
| NPL | 6785 | 29.61 | 19.99 (9.05, 44.28) |
| SDN | 481 | 27.71 | 19.68 (9.23, 43.61) |
| IDN | 3979 | 25.48 | 19.31 (9.06, 44.34) |
| CMR | 2012 | 29.58 | 19.04 (9.16, 44.05) |
| VNM | 36270 | 31.18 | 18.04 (9.00, 44.45) |
| PHL | 87859 | 35.34 | 17.72 (8.95, 44.53) |
| ECU | 7169 | 27.24 | 16.33 (9.19, 43.89) |
| PER | 5307 | 32.17 | 16.27 (9.24, 43.73) |
| LAO | 196 | 26.99 | 16.04 (9.52, 41.59) |

| Countries/regions | Modelled population size (n) | Mean entry age (years) | Estimated LTBI prevalence % (95% CI) |
|---|---|---|---|
| BGD | 14068 | 28.08 | 15.92 (9.13, 44.09) |
| HTI | 5618 | 24.02 | 15.89 (9.15, 43.97) |
| GTM | 87555 | 19.13 | 15.74 (8.94, 44.53) |
| NGA | 19451 | 27.88 | 15.54 (9.13, 44.12) |
| HND | 126561 | 18.61 | 15.32 (8.92, 44.57) |
| THA | 9589 | 26.88 | 15.24 (9.16, 43.91) |
| MAR | 396 | 41.54 | 15.08 (9.54, 41.29) |
| IND | 202047 | 29.82 | 14.76 (8.85, 44.56) |
| KHM | 786 | 13.60 | 14.56 (9.42, 42.49) |
| PAK | 12346 | 28.18 | 14.39 (9.20, 43.81) |
| BIH | 44 | 24.68 | 14.38 (9.69, 38.25) |
| ZAF | 5812 | 23.49 | 14.15 (9.31, 43.26) |
| BOL | 583 | 16.62 | 13.37 (9.56, 41.46) |
| AFG | 9852 | 15.54 | 13.08 (9.25, 43.60) |
| CHN | 324285 | 33.65 | 13.06 (8.92, 44.54) |
| MYS | 3943 | 15.96 | 12.77 (9.36, 42.99) |
| GHA | 6508 | 24.58 | 12.74 (9.39, 42.65) |
| GUY | 2629 | 31.20 | 12.58 (9.48, 42.16) |
| UZB | 140 | 35.47 | 12.30 (9.63, 40.77) |
| MEX | 475791 | 28.89 | 12.02 (8.90, 44.56) |
| YEM | 1276 | 17.58 | 11.90 (9.52, 41.26) |
| HRV | 37 | 26.03 | 11.75 (9.75, 35.12) |
| SLV | 60253 | 20.18 | 11.60 (9.16, 44.03) |
| DOM | 32730 | 26.21 | 11.48 (9.26, 43.61) |
| LKA | 430 | 26.75 | 11.45 (9.63, 39.99) |
| PAN | 383 | 23.05 | 11.25 (9.69, 36.69) |
| IRQ | 552 | 31.14 | 11.05 (9.64, 39.31) |
| PRT | 129 | 34.17 | 10.99 (9.70, 33.74) |
| ROU | 1692 | 29.62 | 10.89 (9.60, 40.30) |
| HKG | 916 | 28.32 | 10.81 (9.62, 39.13) |
| MDA | 534 | 27.51 | 10.70 (9.69, 36.72) |
| FJI | 174 | 21.59 | 10.58 (9.72, 34.77) |
| NIC | 3756 | 20.40 | 10.54 (9.55, 41.17) |
| RUS | 9152 | 29.95 | 10.51 (9.41, 41.82) |
| UKR | 7895 | 29.25 | 10.51 (9.46, 41.82) |
| KOR | 47193 | 29.31 | 10.47 (9.25, 43.64) |
| ALB | 2096 | 30.72 | 10.30 (9.59, 41.06) |
| SAU | 7607 | 22.66 | 10.08 (9.45, 41.91) |
| TUR | 6811 | 31.54 | 9.95 (9.54, 41.28) |
| COL | 24254 | 28.82 | 9.82 (9.36, 42.76) |
| GRD | 6 | 56.00 | 9.76 (9.55, 22.31) |

| Countries/regions | Modelled population size (n) | Mean entry age (years) | Estimated LTBI prevalence % (95% CI) |
|---|---|---|---|
| LTU | 167 | 24.35 | 9.69 (9.56, 32.98) |
| CPV | 26 | 2.00 | 9.67 (9.74, 32.55) |
| TTO | 777 | 30.12 | 9.53 (9.70, 37.42) |
| BLZ | 15 | 14.20 | 9.44 (9.51, 23.49) |
| URY | 82 | 23.11 | 9.29 (9.58, 28.69) |
| BRA | 52514 | 27.49 | 9.28 (9.29, 43.32) |
| GRC | 31 | 47.00 | 9.06 (9.75, 32.13) |
| BGR | 284 | 37.17 | 8.92 (9.75, 33.10) |
| IRN | 4922 | 33.25 | 8.79 (9.52, 41.22) |
| EGY | 4368 | 27.54 | 8.78 (9.64, 39.25) |
| ARG | 2051 | 25.08 | 8.64 (9.69, 37.84) |
| POL | 1871 | 28.73 | 8.63 (9.68, 37.77) |
| ARM | 401 | 18.85 | 8.46 (9.74, 34.80) |
| TWN | 11775 | 32.21 | 8.33 (9.55, 41.01) |
| SYR | 482 | 17.85 | 8.15 (9.69, 38.34) |
| VEN | 48089 | 29.90 | 7.93 (9.40, 42.51) |
| ITA | 5548 | 33.67 | 7.91 (9.68, 37.71) |
| BEL | 384 | 25.40 | 7.90 (9.69, 29.45) |
| CRI | 869 | 29.22 | 7.79 (9.74, 34.85) |
| AUT | 436 | 22.91 | 7.69 (9.71, 34.45) |
| JOR | 2713 | 18.95 | 7.59 (9.71, 36.92) |
| CUB | 24270 | 32.22 | 7.56 (9.53, 41.06) |
| ESP | 9654 | 24.24 | 7.50 (9.63, 39.80) |
| BLR | 610 | 26.92 | 7.42 (9.67, 34.72) |
| HUN | 237 | 31.41 | 7.32 (9.40, 25.82) |
| JAM | 14939 | 26.99 | 7.23 (9.62, 40.08) |
| LBN | 1215 | 30.12 | 7.06 (9.70, 31.55) |
| FRA | 18842 | 29.56 | 6.96 (9.57, 40.71) |
| CHL | 3595 | 29.81 | 6.81 (9.71, 36.08) |
| CZE | 2503 | 22.07 | 6.80 (9.62, 38.84) |
| BRB | 14 | 27.50 | 6.73 (7.36, 16.64) |
| JPN | 36544 | 27.01 | 5.84 (9.64, 39.24) |
| ISR | 1772 | 26.41 | 5.81 (9.71, 30.65) |
| IRL | 969 | 27.12 | 5.78 (9.62, 29.08) |
| SWE | 1263 | 26.74 | 5.59 (9.66, 27.52) |
| CHE | 1085 | 23.20 | 5.56 (9.52, 28.43) |
| GBR | 20621 | 33.27 | 5.51 (9.71, 35.33) |
| DEU | 16244 | 26.23 | 5.28 (9.70, 35.39) |
| CAN | 63390 | 46.14 | 5.11 (9.65, 37.92) |
| NLD | 1353 | 25.43 | 5.10 (9.51, 25.03) |
| AUS | 7198 | 26.90 | 4.48 (9.63, 29.09) |

#### Appendix 2

##### Clinical flowchart of the post-arrival screening strategies

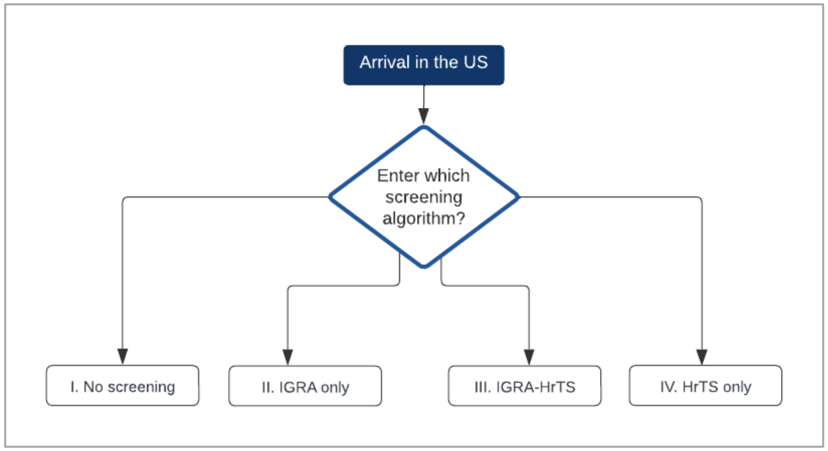

**Figure E1-1. Four post-arrival screening strategies.**

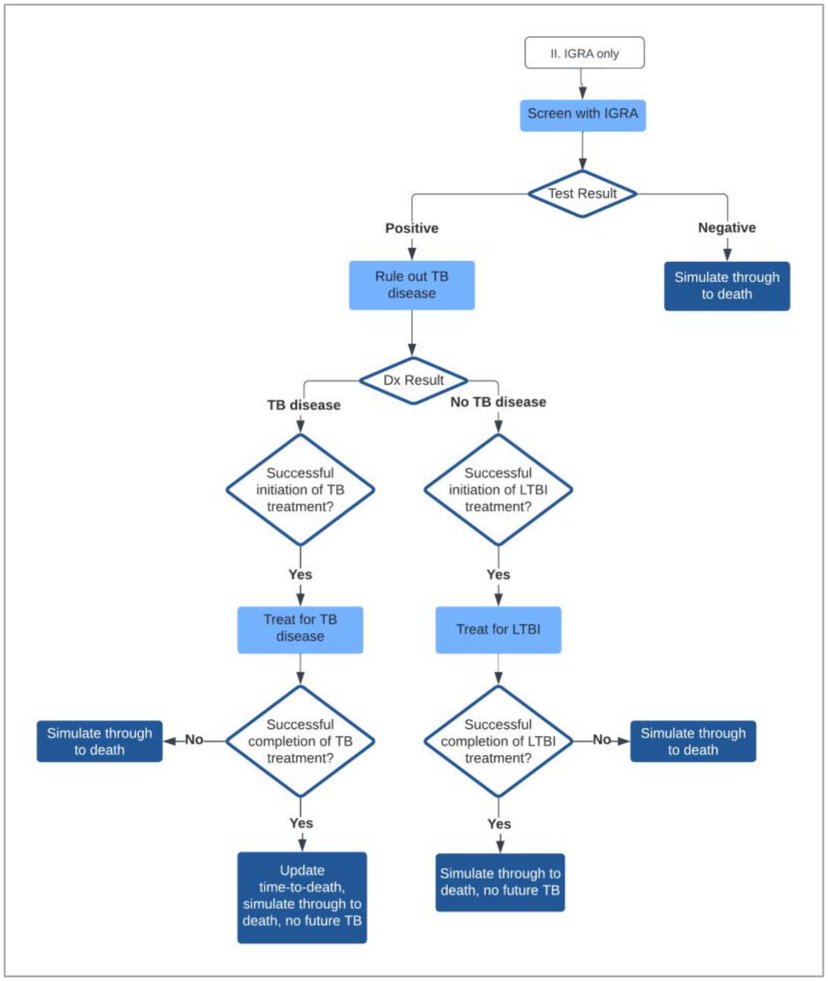

**Figure E1-2. Strategy II IGRA only.**

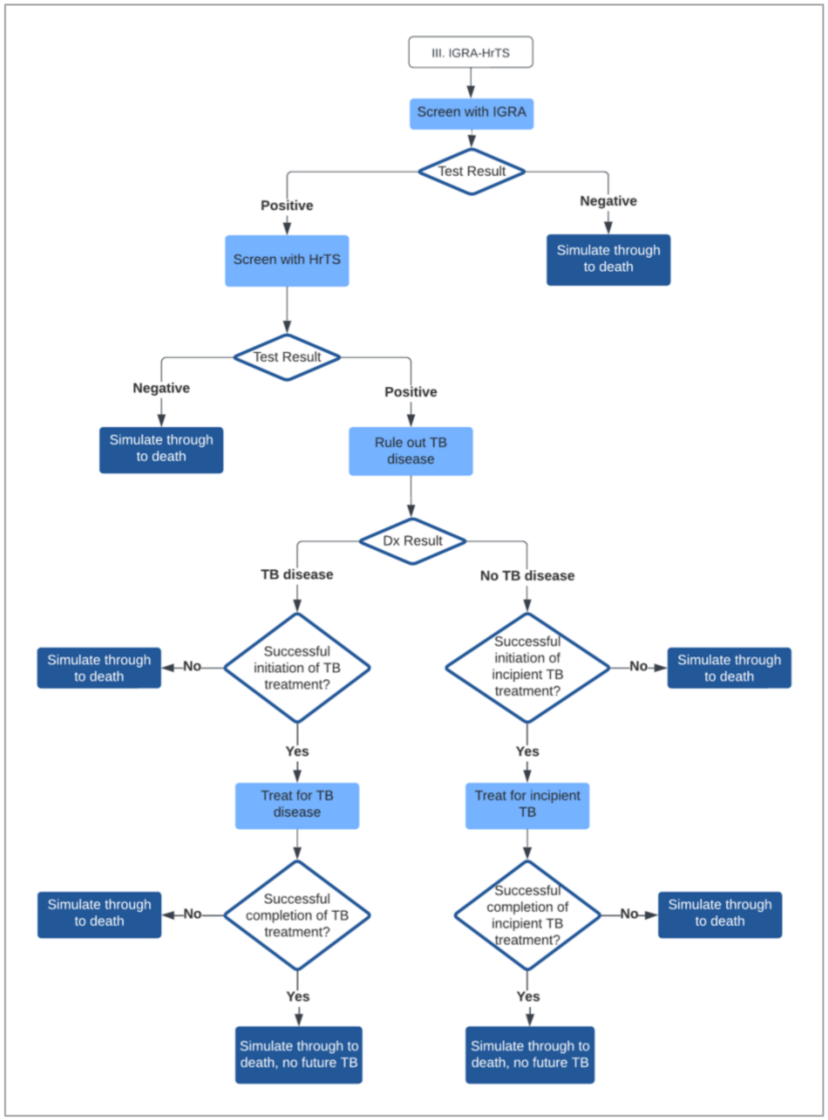

**Figure E1-3. Strategy III IGRA-HrTS.**

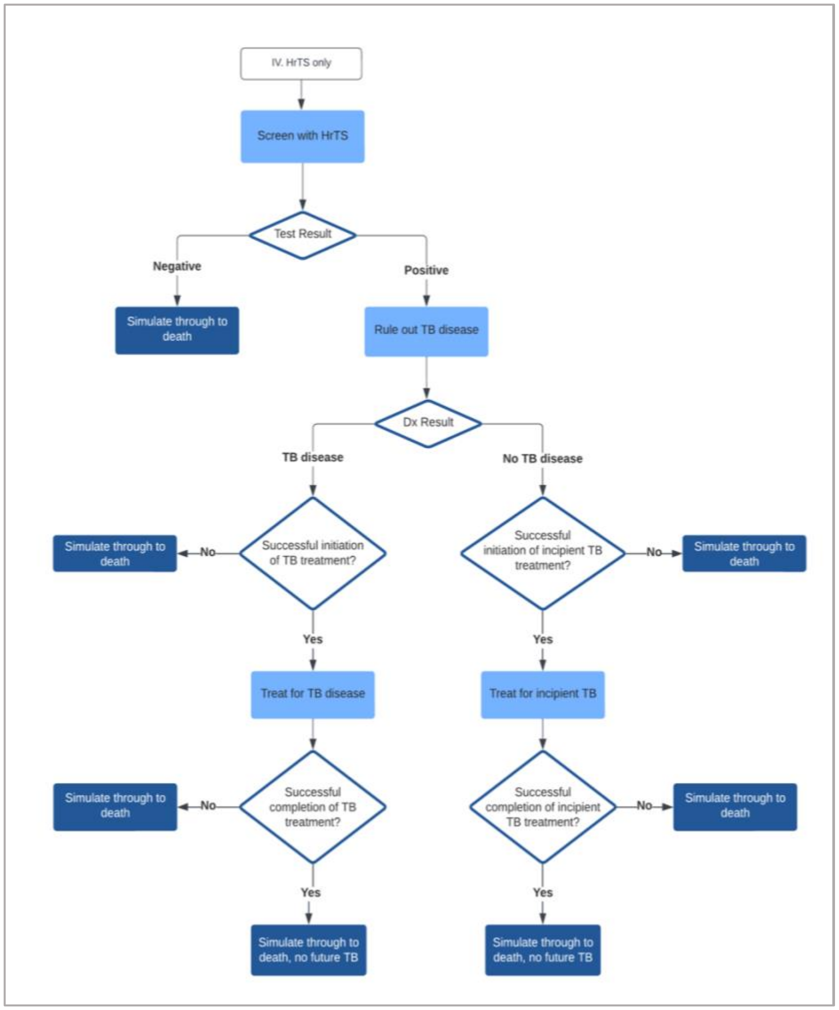

**Figure E1-4. Strategy IV HrTS only.**

#### Appendix 3

##### Estimation of the LTBI prevalence of the study cohort

Prevalence of *Mtb* infection among migrants in 2019 by age and country-of-origin was estimated as follows. We first used the LTBI prevalence data reported in Collins *et al.* and averaged country-of-origin specific TB incidence data from reported in Hill *et al.*[1,2] to train an ordinary least squares regression that minimizes the sum of squared error:

$$\min \sum_{i=1}^{23} \left( \log(Y_i) - \log\left( \frac{1}{1 + e^{-\beta_0 + \beta_1 X_i}} \right) \right)^2$$

where $Y_i$ is country-of-origin specific LTBI prevalence among migrants and $X_i$ is the log of country-of-origin specific TB incidence among migrants. We had both LTBI prevalence estimates and TB incidence data for a total of 23 countries.

We then used this model to predict the LTBI prevalence (%) among the 2019 entry cohort by age and country of origin and its associated 95% confidence interval, based on TB incidence rates obtained from the TB risk model by Hill *et al*.[1] Lastly, we calibrated the LTBI prevalence estimates such that the overall LTBI prevalence among the 2019 entry cohort matched the most recent national level LTBI prevalence estimate (12.4%) among non-US born population reported in NHANES.[3] Without the calibration step, the model estimated prevalence would be overestimated as the data reported in Collins *et al.* were for higher-risk populations.

#### Appendix 4

##### Discrete event simulation (DES) model

*Time-to-event distributions.* At the start of the simulation period, each individual was assigned a time-to-TB value. This value was sampled from empirical survival functions of TB disease based on migrants' entry age, country-of-origin, year-of-entry, and time-since-entry to the US. The survival functions were derived from the annual TB risk functions, which were estimated using the 2019 cohort demographic data and the fitted TB risk prediction model developed in *Hill et al.*[1] At the beginning of the simulation period, each individual was also assigned a time-to-death value, which was sampled from survival functions for the general population, by starting age. The age-specific rates were informed by the male and female 2017 US Life Tables for foreign born individuals,[4] assuming 50% male and 50% female. Both time-to-TB and time-to-death were modelled in days. We used the `approx()` function in R to convert the time scale from year to day using linear interpolation.[5]

*Modelling TB disease.* TB cases not identified through the screening strategies were assumed to be identified when individuals sought healthcare for TB symptoms. Those identified with symptomatic disease were assumed to experience a higher case fatality rate, as compared to individuals diagnosed with TB via the screening (Appendix 5 Table 1). The relative mortality rates for those with TB disease were calibrated using NTSS treatment outcome data, which indicated that 1.6% of TB cases died at TB diagnosis and 5.4% of those who were alive at TB diagnosis died before treatment completion (unpublished CDC treatment outcome data for TB cases among non-US born individuals between 2010-2019, provided by Julie Self, Lauren Lambert, and Bob Pratt from the U.S. CDC Surveillance Team, Division of Tuberculosis Elimination, 2022-10-06). Individuals surviving TB were assumed to experience an increased risk (RR = 1.78) for six years,[6] after which the mortality rate returned to background mortality (RR=1).

*Modelling treatment for TB disease and incipient TB.* We assumed individuals diagnosed with incipient TB understand that they are at a higher risk of falling ill and consequently have a higher treatment initiation rate compared to those diagnosed with LTBI (88.1% vs 76.2%). For the same reason, we assumed the treatment completion rate for incipient TB is higher than that for LTBI (92.7% vs 90.3%). In our model, we assumed those treated for LTBI had a 64% chance of clearing TB infection, and that the incipient TB treatment had the same efficacy as TB disease treatment.

*Sampling*. Due to constraints in computational resources, instead of simulating individual trajectories for all migrants in the study cohort, we simulated the trajectories for 50% of the migrant cohort from the Philippines and 10% of the migrant cohort from China, India, and Mexico. The sampled cohorts were representative of their parent cohorts. Their simulated results were proportionally upweighted in outcome calculations.

### Appendix 5

#### Parameter tables

##### Table E2. Parameter table

| Parameters | Mean | *Source and comments* |
|---|---|---|
| Background mortality rate | Age-specific | 2017 US Life Tables for foreign born individuals,[4] see Appendix 4. |
| **Entering post-arrival screening program** | | |
| Time to post-arrival screening (month) | 1 | (Model setting) |
| **Testing related assumptions** | | |
| % Screened with IGRA among those identified to be screened | 100 | (Model setting) |
| % Screened for TB among IGRA positives (Strategy II IGRA-TB) | 100 | (Model setting) |
| % Screened with RNA test among IGRA positives (Strategy III IGRA-RNA-TB) | 100 | (Model setting) |
| % Screened for TB among RNA positives (Strategy III IGRA-RNA-TB) | 100 | (Model setting) |
| P (tested negative for IGRA \| no TB infection) | 0.985 | Stout 2018 *Thorax*[7] |
| P (tested positive for IGRA \| TB infection) | 0.789 | Stout 2018 *Thorax*[7] |
| P (diagnosed to not have TB disease \| Healthy) | 1.00 | Assumption |
| P (diagnosed to not have TB disease \| LTBI) | 1.00 | Assumption |
| P (diagnosed with TB disease \| TB) | 1.00 | Assumption |
| P (tested negative for the signature test \| no TB infection) | 0.90 | WHO TPP[8] |
| P (tested positive for the signature test \| TB) | 0.90 | WHO TPP[8] |
| P (tested positive for the signature test \| LTBI, time-to-TB <= 2 years) | 0.90 | WHO TPP[8] |
| P (tested positive for the signature test \| LTBI, time-to-TB > 2 years) | 0.10 | Assumption, the test cannot distinguish cases from non-cases of future TB disease occurring > 2 years from time of testing |
| **Treatment related assumptions** | | |
| ***Mtb* infection** ***(Treatment: once-weekly isoniazid-rifapentine for 12 weeks (3HP))*** | | |
| P (Initiated treatment for *Mtb* infection \| diagnosed as *Mtb* infection) | 0.762 | CDC State and City TB Report 2020[9] |

| Parameters | Mean | *Source and comments* |
|---|---|---|
| P (Completed treatment for *Mtb* infection \| successfully initiated *Mtb* infection treatment) | 0.903 | Sandul 2017 *CID*[10] |
| Treatment length among those who completed *Mtb* infection treatment (weeks)<br>Treatment length among those who did not complete *Mtb* infection treatment (weeks) | 12<br>3 | 3HP<br>Sterling 2015 *CID*[11] |
| P (Cured of TB infection \| did not complete *Mtb* infection treatment)<br>P (Cured of TB infection \| completed *Mtb* infection treatment) | 0<br>0.64 | Assumption<br>Zenner 2017 *Annal Int Med*[12] |
| Effect of *Mtb* infection treatment among those cured of *Mtb* infection | Restore utility weight to 1; will never develop TB disease in lifetime | |
| ***Incipient TB***<br>***(Treatment: 1-month initial phase of daily isoniazid- rifampicin, followed by 3-month continuation phase of isoniazid-rifampicin (*1HR daily, 3HR thrice weekly*))*** | | |
| P (Initiated treatment for incipient TB \| diagnosed as incipient TB) | 0.881 | Assumption, arithmetic mean of initiation rate for TB and LTBI treatments, with 95% CrI (0.76, 0.95) |
| P (Completed treatment for incipient TB \| successfully initiated incipient TB treatment) | 0.916 | Assumption, same as TB treatment completion rate, but greater variance, with 95% CrI (0.88, 0.97) |
| Treatment length among those who completed incipient TB treatment (weeks)<br>Treatment length among those who did not complete incipient TB treatment (weeks) | 18<br>3 | 1HR daily + 3HR thrice weekly, Hamada 2016 *Ann Am Thorac Soc*[13]<br>Assumption, same as LTBI treatment |
| P (Cured of TB infection \| did not complete incipient TB Treatment)<br>P (Cured of TB infection \| completed incipient TB Treatment) | 0<br>0.965 | Assumption<br>Assumption, same as TB treatment |
| Effect of incipient TB treatment among those cured of incipient TB | Restore utility weight to 1; will never develop TB disease in lifetime | |

| Parameters | Mean | *Source and comments* |
|---|---|---|
| ***TB disease*** ***(Treatment: 2-month initial phase of daily or thrice weekly rifampin, isoniazid, pyrazinamide, ethambutol daily, followed by 4-month continuation phase of isoniazid- rifampin (2RIPE + 4HP)*** | | |
| P (Initiated treatment for TB disease \| diagnosed with TB disease, via active screening or through passive detection) | 1 | Assumption |
| Time between symptom onset and diagnosis for TB cases not identified through the post-arrival screening program (year) | 0.5 | Assumption, with 95%CrI (0.25, 0.74), about 3-9 months |
| P (Dead at TB diagnosis and therefore not eligible to begin TB treatment) | 0.016 | Unpublished CDC data, provided by Julie Self, Lauren Lambert, and Bob Pratt from the U.S. CDC Surveillance Team, Division of Tuberculosis Elimination, 2022-10-06. |
| P (Completed treatment for TB \| did not die on TB treatment) | 0.916 | Unpublished CDC data, provided by Julie Self, Lauren Lambert, and Bob Pratt from the U.S. CDC Surveillance Team, Division of Tuberculosis Elimination, 2022-10-06. |
| P (Died on TB treatment \| alive at TB diagnosis) | 0.0544 | Unpublished CDC data, provided by Julie Self, Lauren Lambert, and Bob Pratt from the U.S. CDC Surveillance Team, Division of Tuberculosis Elimination, 2022-10-06. |
| Treatment length among those who completed TB treatment (weeks)<br>Treatment length among those who did not complete TB treatment(weeks) | 26<br>3 | 2 RIPE + 4HP<br>Assumption, same as *Mtb* infection treatment |
| Time between TB diagnosis and treatment initiation (days) | 3 | Assumption |
| Time between repeated treatments after default (months) | 6 | Assumption |
| P (Cured of *Mtb* infection \| did not complete TB Treatment)<br>P (Cured of *Mtb* infection \| completed TB Treatment) | 0<br>1 | Assumption<br>Assumption |

| Parameters | Mean | *Source and comments* |
|---|---|---|
| Effect of TB treatment among those cured of TB Disease | Restore QALY to 1, will never develop TB disease in lifetime, but continue to experience elevated mortality risk for 6 years after completion of TB treatment (HR = 1.78) and will experience the background mortality following that | Lee Rodriguez 2020 *JAMA Netw Open*[6] |
| **Economic Evaluation** | | |
| Utility weight for no *Mtb* infection | 1 | |
| Utility weight for *Mtb* infection and incipient TB not on treatment | 1 | |
| Utility weight for TB not on treatment or on *Mtb* infection or incipient TB treatment | 0.76 | Guo 2009 *Health Qual Life Outcomes,*[14] HUI3 estimates |
| Utility weight for *Mtb* infection and incipient TB on *Mtb* infection treatment | 0.998 | Jo 2021 *Clin Infect Dis Off Publ Infect Dis Soc Am* [15] |
| Utility weight for *Mtb* infection and incipient TB on incipient TB Treatment | 0.998 | Following Jo 2019 *Clin Infect Dis Off Publ Infect Dis Soc Am*,[15] 0.005*0.75 + (1-0.005)*1 = 0.998 |
| Utility weight for anyone on TB treatment | 0.92 | Guo 2009 *Health Qual Life Outcomes*,[14] HUI3 estimates for well-controlled TB |
| Utility weight for anyone after TB treatment completion if cured | 0.99 | Menzies 2021 *Lancet GH* [16] |
| Cost of *Mtb* infection treatment for those successfully initiated treatment (same for cases of treatment completion and default) (3HP) (US dollars) | 520 | CDC Website[17] |

| Parameters | Mean | *Source and comments* |
|---|---|---|
| Cost of incipient TB treatment for those successfully initiated treatment (same for cases of treatment completion and default) (1HR daily, 3HR thrice weekly) (US dollars) | 880 | Calculated based on numbers on the CDC Website<br>463 (1HR daily, 90 doses) +<br>(463 – 355) + (355 – 76) + 76 *12/30 (3HR, thrice weekly) = 880.4 |
| Cost of TB treatment for those successfully initiated treatment (same for cases of treatment completion and default) (US dollars) | 24,661 | CDC Website[18] |
| Cost of IGRA (2021 US dollars)<br>Cost of HrTS (2021 US dollars)<br>Cost of TB screening (Chest x-ray 2 views) (2021 US dollars) | 61.69<br>30<br>33.2 | CMS Clinical Lab Fee Schedule 2020[19]<br>Assumption<br>CMS Chest X-Ray Policy |
| Annual non-TB health care costs (2021 US dollars) | Varied by age | Jiao 2021 *Value Health* [21] |
| Annual non-health care expenditures (2021 US dollars) | Varied by age category | BLS Consumer Expenditure Surveys 2021[22] |
| Annual total (market and non-market) labor productivity in the U.S. (2021 US dollars) | Varied by age | Grosse 2016 *J Med Econ*[23] |
| Annual discount rate | 0.03 | Neumann 2016 Cost-Effectiveness in Health and Medicine[24] |

##### Table E3. Distributional characteristics of the uncertain input parameters for probabilistic sensitivity analysis

| Parameters | Mean | Median | 95% Credible Interval | Uncertainty Distribution | *Source and comments* |
|---|---|---|---|---|---|
| Time between symptom onset and diagnosis for TB cases not identified through the post-arrival screening program (year) | 0.5 | 0.50 | 0.26 – 0.74 | Lognormal (0.5, 0.125) | Assumption, with 95%CrI (0.25, 0.74), about 3-9 months |
| P (tested negative for IGRA \| no *Mtb* infection) | 0.985 | 0.987 | 0.961 – 0.998 | Beta (169.11, 2.58) | Stout 2018 *Thorax*[7] |

| Parameters | Mean | Median | 95% Credible Interval | Uncertainty Distribution | *Source and comments* |
|---|---|---|---|---|---|
| P (tested positive for IGRA \| *Mtb* infection) | 0.789 | 0.792 | 0.696 – 0.902 | Beta (48.74, 13.03) | Stout 2018 *Thorax*[7] |
| P (Initiated treatment for *Mtb* infection \| diagnosed as *Mtb* infection) | 0.762 | 0.762 | 0.750 – 0.773 | Beta (4235, 1325) | CDC State and City TB Report 2020 |
| P (Completed treatment for *Mtb* infection \| successfully initiated *Mtb* infection treatment) | 0.903 | 0.902 | 0.886 – 0.919 | Beta (1168, 126) | Sandul 2017 *CID*[10] |
| P (Cured of *Mtb* infection \| completed *Mtb* infection treatment) | 0.64 | 0.65 | 0.27 – 0.82 | Beta (7.16, 4.03) | Zenner 2017 *Annal Int Med*[12] |
| P (Initiated treatment for incipient TB \| diagnosed as incipient TB) | 0.881 | 0.887 | 0.76 – 0.95 | Beta (40.06, 5.41) | Assumption, arithmetic mean of initiation rate for TB and LTBI treatments, with 95% CrI (0.76, 0.95) |
| P (Completed treatment for incipient TB \| successfully initiated incipient TB treatment) | 0.916 | 0.919 | 0.86 – 0.95 | Beta (122.99, 9.68) | Assumption, same as TB treatment completion rate, but greater variance, with 95% CrI (0.88, 0.97) |
| P (Cured of *Mtb* infection \| completed incipient TB Treatment) | 0.64 | 0.65 | 0.27 – 0.82 | Beta (7.16, 4.03) | Assumption, same as LTBI treatment cure rate |
| P (Dead at TB diagnosis and therefore not eligible to begin TB treatment) | 0.016 | 0.016 | 0.014 – 0.018 | Beta (0.387, 23.80) | Unpublished CDC data, provided by Julie Self, Lauren Lambert, and Bob Pratt from the U.S. CDC Surveillance Team, Division of Tuberculosis Elimination, 2022- |

| Parameters | Mean | Median | 95% Credible Interval | Uncertainty Distribution | *Source and comments* |
|---|---|---|---|---|---|
| | | | | | 10-06, with 95% CrI (0.01, 0.02) |
| P (Died on TB treatment \| alive at TB diagnosis) | 0.0544 | 0.0541 | 0.035 – 0.074 | Beta (27.92, 485.48) | Unpublished CDC data, provided by Julie Self, Lauren Lambert, and Bob Pratt from the U.S. CDC Surveillance Team, Division of Tuberculosis Elimination, 2022-10-06, with 95% CrI (0.035, 0.075) |
| P (Completed treatment for TB \| did not die on TB treatment) | 0.916 | 0.917 | 0.90 – 0.94 | Beta (450.16, 41.28) | Unpublished CDC data, provided by Julie Self, Lauren Lambert, and Bob Pratt from the U.S. CDC Surveillance Team, Division of Tuberculosis Elimination, 2022-10-06, with 95% CrI (0.90, 0.95) |
| Cost of IGRA (2021 US dollars) | 64.38 | 63.10 | 49.10 – 79.96 | Gamma (59.68, 1.08) | Assumption, range = [50, 100] |
| Cost of TB screening (Chest x-ray 2 views) (2021 US dollars) | 34.49 | 34.31 | 25.26 – 45.55 | Gamma (47.58, 0.72) | Assumption, range = [20, 50] |
| Cost of *Mtb* infection treatment for those successfully initiated treatment (same for cases of treatment completion and default) (3HP) – healthcare costs (2021 US dollars) | 427.98 | 429.56 | 221.69 – 705.40 | Gamma (13.46, 31.81) | Assumption, range = [300, 1000] |
| Cost of *Mtb* infection treatment for those successfully initiated treatment (same for cases of treatment completion and | 112.18 | 111.88 | 93.43 – 131.83 | Gamma (125.84, 0.89) | Assumption, Range = [80, 140] |

| Parameters | Mean | Median | 95% Credible Interval | Uncertainty Distribution | *Source and comments* |
|---|---|---|---|---|---|
| default) (3HP) – non-healthcare costs (2021 US dollars) | | | | | |
| Cost of incipient TB treatment for those successfully initiated treatment (same for cases of treatment completion and default) (1HR daily, 3HR thrice weekly) – healthcare costs (2021 US dollars) | 513.92 | 501.72 | 281.45 – 828.35 | Gamma (14.86, 34.59) | Assumption, range = [400, 1200] |
| Cost of incipient TB treatment for those successfully initiated treatment (same for cases of treatment completion and default) (1HR daily, 3HR thrice weekly) – non-healthcare costs (2021 US dollars) | 149.57 | 149.08 | 125.81 – 176.09 | Gamma (125.84, 1.19) | Assumption, Range = [110, 190] |
| Utility weight for TB not on treatment or on LTBI or incipient TB treatment | 0.76 | 0.76 | 0.70 – 0.82 | 1 – Gamma (64, 0.00375) | Guo 2009 Health Qual Life Outcomes,[14] HUI3 estimates |
| Utility weight for anyone on TB treatment | 0.91 | 0.94 | 0.71 – 0.99 | 1 – Gamma (1.31, 0.061) | Guo 2009 Health Qual Life Outcomes,[14] HUI3 estimates for well-controlled TB |
| Utility weight for anyone after TB treatment completion if cured | 0.99 | 0.99 | 0.98 – 0.99 | 1 – Gamma (16, 0.00075) | Menzies 2021 *Lancet GH*[16] |
| Cost of TB treatment for those successfully initiated treatment (same for cases of treatment completion and default) – healthcare costs (2021 US dollars) | 20,993.27 | 20811.99 | 16510.80 – 26347.69 | Gamma (70.51, 297.71) | Assumption, range = [15,000, 30,000] |
| Cost of TB treatment for those successfully initiated treatment (same for cases of treatment completion and default) – non-healthcare costs (2021 US dollars) | 4622.24 | 4647.67 | 3900.13 – 5459.68 | Gamma (145.40, 31.79) | Assumption, Range = [3500, 5800] |

### Appendix 6

#### Screening and treatment cascade by strategy and risk category

##### Table E4. Expected percentage of population screened, by test type

| Risk Category[1] | Percent of population screened, by test type (%, 95% CI) | | | | | | |
|---|---|---|---|---|---|---|---|
| | Strategy II. IGRA only | | Strategy III. IGRA-HrTS | | | Strategy IV. HrTS only | |
| | IGRA | TB diagnosis | IGRA | HrTS | TB diagnosis | HrTS | TB diagnosis |
| **Whole cohort** | 100 | 11.1 (9.2 – 13.4) | 100 | 11.1 (9.2 – 13.4) | 1.1 (0.9 – 1.4) | 100 | 10.0 (10.0 – 10.0) |
| **I** | 100 | 14.8 (12.6 – 17.3) | 100 | 14.8 (12.6 – 17.3) | 1.6 (1.3 – 1.8) | 100 | 10.2 (10.2 – 10.3) |
| **II** | 99.9 | 13.5 (11.3 – 15.8) | 99.9 | 13.5 (11.3 – 15.8) | 1.4 (1.2 – 1.7) | 99.9 | 10.1 (10.1 – 10.2) |
| **III** | 100 | 10.9 (9.1 – 13.2) | 100 | 10.9 (9.1 – 13.2) | 1.1 (0.9 – 1.4) | 100 | 10.0 (10.0 – 10.0) |
| **IV** | 100 | 6.3 (4.8 – 8.6) | 100 | 6.3 (4.8 – 8.6) | 0.6 (0.5 – 0.9) | 100 | 9.8 (9.8 – 9.9) |

[1] *Epidemiological categorization of migrant populations based on TB incidence per 100k in 2019 for their country-of-origin:* risk category I (≥300); risk category II (100-300), risk category III (10-100), risk category IV (0-10).

#### Table E5. Expected percentage of population treated, by regimen

| Risk Category[1] | Percent of population treated, by regimen (%, 95% CI) | | | | | | |
|---|---|---|---|---|---|---|---|
| | Strategy I. No testing | Strategy II. IGRA only | | Strategy III. IGRA-HrTS | | Strategy IV. HrTS only | |
| | TB disease | *Mtb* infection | TB disease | Incipient TB | TB disease | Incipient TB | TB disease |
| **Whole cohort** | 0.3 (0.2 – 0.3) | 8.5 (7.1 – 10.3) | 0.2 (0.1 – 0.2) | 1.0 (0.8– 1.2) | 0.2 (0.2 – 0.3) | 8.8 (7.8 – 9.6) | 0.2 (0.2 – 0.3) |
| **I** | 0.9 (0.8 – 1.0) | 11.3 (9.6 – 13.1) | 0.6 (0.5 – 0.8) | 1.4 (1.1 – 1.6) | 0.8 (0.7 – 0.9) | 9.0 (8.0 – 9.8) | 0.8 (0.7 – 0.9) |
| **II** | 0.5 (0.4 – 0.6) | 10.3 (8.7 – 12.1) | 0.3 (0.2 – 0.4) | 1.3 (1.0 – 1.5) | 0.5 (0.4 – 0.5) | 8.9 (7.9 – 9.7) | 0.5 (0.4 – 0.5) |
| **III** | 0.2 (0.2 – 0.2) | 8.4 (6.9 – 10.1) | 0.1 (0.1 – 0.2) | 1.0 (0.8 – 1.2) | 0.2 (0.2 – 0.2) | 8.8 (7.8 – 9.5) | 0.2 (0.2 – 0.2) |
| **IV** | 0.02 (0.01 – 0.02) | 4.8 (3.7 - 6.6) | 0.01 (0.01 – 0.02) | 0.6 (0.4 – 0.8) | 0.01 (0.01 – 0.02) | 8.6 (7.6 – 9.4) | 0.01 (0.01 – 0.02) |

[1] *Epidemiological categorization of migrant populations based on TB incidence per 100k in 2019 for their country-of-origin:* risk category I (≥300); risk category II (100-300), risk category III (10-100), risk category IV (0-10).

### Appendix 7

#### Cost-Effectiveness Analysis Impact Inventory

#### Table E6. Impact inventory

| Sector | Type of impact<br>Categories impacted within each sector with unit of measure if relevant in the Analysis | Included in the Analysis from this perspective?<br>Healthcare sector | Societal |
|---|---|---|---|
| FORMAL HEALTHCARE SECTOR | | | |
| **Health** | Health outcomes (effects): | | |
| | Longevity effects, Years | ✓ | ✓ |
| | Health-related quality-of-life effects, QALYs | ✓ | ✓ |
| | Medical costs: | | |
| | TB screening related costs (payers and patients) | ✓ | ✓ |
| | Future TB related medical costs (payers and patients) | ✓ | ✓ |
| | Future unrelated medical costs (payers and patients) | ✓ | ✓ |
| INFORMAL HEALTHCARE SECTOR | | | |
| **Health** | Patient time costs | NA | ✓ |
| | Unpaid caregiver time costs | NA | □ |
| | Transportation costs | NA | □ |
| NON-HEALTHCARE SECTOR | | | |
| **Productivity** | Labor market and non-market earnings lost | NA | ✓ |
| **Consumption** | Future consumption unrelated to health, $ | NA | ✓ |
| **Social services** | None | NA | - |
| **Legal/ criminal justice** | None | NA | - |
| **Education** | None | NA | - |

| Sector | Type of impact | Included in the Analysis from this perspective? | |
|---|---|---|---|
| | Categories impacted within each sector with unit of measure if relevant in the Analysis | Healthcare sector | Societal |
| **Housing** | None | NA | - |
| **Environment** | None | NA | - |

***Notes on sources of evidence.*** Please refer to Appendix 5 for relevant parameter input values.

#### Costing methods

For TB-related and TB-unrelated healthcare expenditures, we used the PCE-Health (personal consumption expenditure for health) price indices to express all costs in 2020 US dollars (Table 3 Column 1 on the Agency for Healthcare Research and Quality website here). And because PCE-health has a two-year lag-time, we used the PCE price indices in 2020 and 2021 to bring the numbers to 2021 US dollars (Table 2 Column 3).[24] For non-healthcare expenditures and productivity, we used the PCE price indices (Table 2 Column 3) to express everything in 2021 US dollars.

#### Appendix 8

##### Additional economic evaluation results

**Table E7. Expected per-person gain in quality-adjusted life years (QALY) relative to Strategy I, by risk category.**

| Risk category[1] | Population size | Expected per-person incremental gain in QALY (95% CI) | | |
|---|---|---|---|---|
| | | Strategy II. IGRA-only | Strategy III. IGRA-HrTS | Strategy IV. HrTS-only |
| **I** | 100,778 | 0.00404 (0.00275, 0.00544) | 0.00271 (0.00169, 0.00378) | 0.00282 (0.00168, 0.00396) |
| **II** | 340,454 | 0.00273 (0.00183, 0.00368) | 0.00161 (0.00093, 0.00238) | 0.00153 (0.00077, 0.00237) |
| **III** | 1,409,531 | 0.00098 (0.00063, 0.00135) | 0.00069 (0.00039, 0.00103) | 0.00064 (0.00032, 0.00100) |
| **IV** | 191,462 | 0.00008 (0.00003, 0.00012) | 0.00008 (0.00004, 0.00012) | 0.00001 (-0.00006, 0.00006) |

[1] *Epidemiological categorization of migrant populations based on TB incidence per 100k in 2019 for their country-of-origin:* risk category I (≥300); risk category II (100-300), risk category III (10-100), risk category IV (0-10).

**Table E8. Expected per-person additional cost relative to Strategy I, by risk category.**

| Risk category[1] | Population size | Expected per-person incremental cost (95% CI) | | |
|---|---|---|---|---|
| | | Strategy II. IGRA-only | Strategy III. IGRA-HrTS | Strategy IV. HrTS-only |
| **Healthcare Sector Perspective** | | | | |
| I | 100,778 | 105.9 (74.8, 142.6) | 74.4 (57.3, 93.7) | 73.4 (50.7, 101.2) |
| II | 340,454 | 102.5 (73.2, 137.5) | 73.7 (57.1, 92.1) | 75.2 (53.7, 102.4) |
| III | 1,409,531 | 97.9 (73.4, 127.3) | 72.4 (56.6, 90.5) | 77.1 (57.2, 103.4) |
| IV | 191,462 | 86.9 (68.2, 109.1) | 69.4 (53.8, 86.5) | 77.7 (58.0, 103.2) |
| **Societal Perspective** | | | | |
| I | 100,778 | 139.6 (107.6, 178.3) | 82.2 (65.2, 101.0) | 92.1 (69.3, 121.2) |
| II | 340,454 | 129.4 (99.2, 166.0) | 79.6 (62.9, 98.8) | 92.7 (70.8, 120.8) |
| III | 1,409,531 | 109.2 (84.9, 139.4) | 72.7 (56.8, 91.1) | 89.0 (68.3, 115.3) |
| IV | 191,462 | 92.0 (72.6, 114.5) | 69.9 (54.3, 87.4) | 90.3 (70.4, 116.5) |

[1] *Epidemiological categorization of migrant populations based on TB incidence per 100k in 2019 for their country-of-origin:* risk category I (≥300); risk category II (100-300), risk category III (10-100), risk category IV (0-10).

**Table E9. Expected per-person incremental net monetary benefit (NMB) relative to Strategy I, by risk category.**

| Risk category[1] | Population size | Expected per-person incremental NMB (95% CI) | | |
|---|---|---|---|---|
| | | Strategy II. IGRA-only | Strategy III. IGRA-HrTS | Strategy IV. HrTS-only |
| **Healthcare Sector Perspective** | | | | |
| I | 100,778 | 500.5 (296.6, 719.1) | 332.5 (179.9, 492.4) | 350.1 (166.0, 530.4) |
| II | 340,454 | 306.7 (154.6, 463.5) | 167.6 (60.2, 284.1) | 153.9 (34.1, 284.1) |
| III | 1,409,531 | 48.7 (-11.7, 110.0) | 31.6 (-17.6, 84.2) | 18.7 (-37.9, 77.9) |
| IV | 191,462 | -75.2 (-99.8, -54.7) | -58.1 (-76.1, -41.8) | -76.4 (-103.4, -54.4) |
| **Societal Perspective** | | | | |
| I | 100,778 | 466.8 (270.1, 679.8) | 324.7 (174.7, 483.0) | 331.5 (149.7, 510.5) |
| II | 340,454 | 279.8 (129.0, 431.6) | 161.6 (54.8, 277.5) | 136.3 (17.9, 267.6) |
| III | 1,409,531 | 37.4 (-23.4, 97.0) | 31.4 (-17.8, 84.2) | 6.8 (-17.8, 66.0) |
| IV | 191,462 | -80.2 (-104.9, -58.8) | -58.6 (-76.7, -41.9) | -89.0 (-117.0, -66.0) |

[1] *Epidemiological categorization of migrant populations based on TB incidence per 100k in 2019 for their country-of-origin:* risk category I (≥300); risk category II (100-300), risk category III (10-100), risk category IV (0-10).

#### Appendix 9

##### Sensitivity analyses

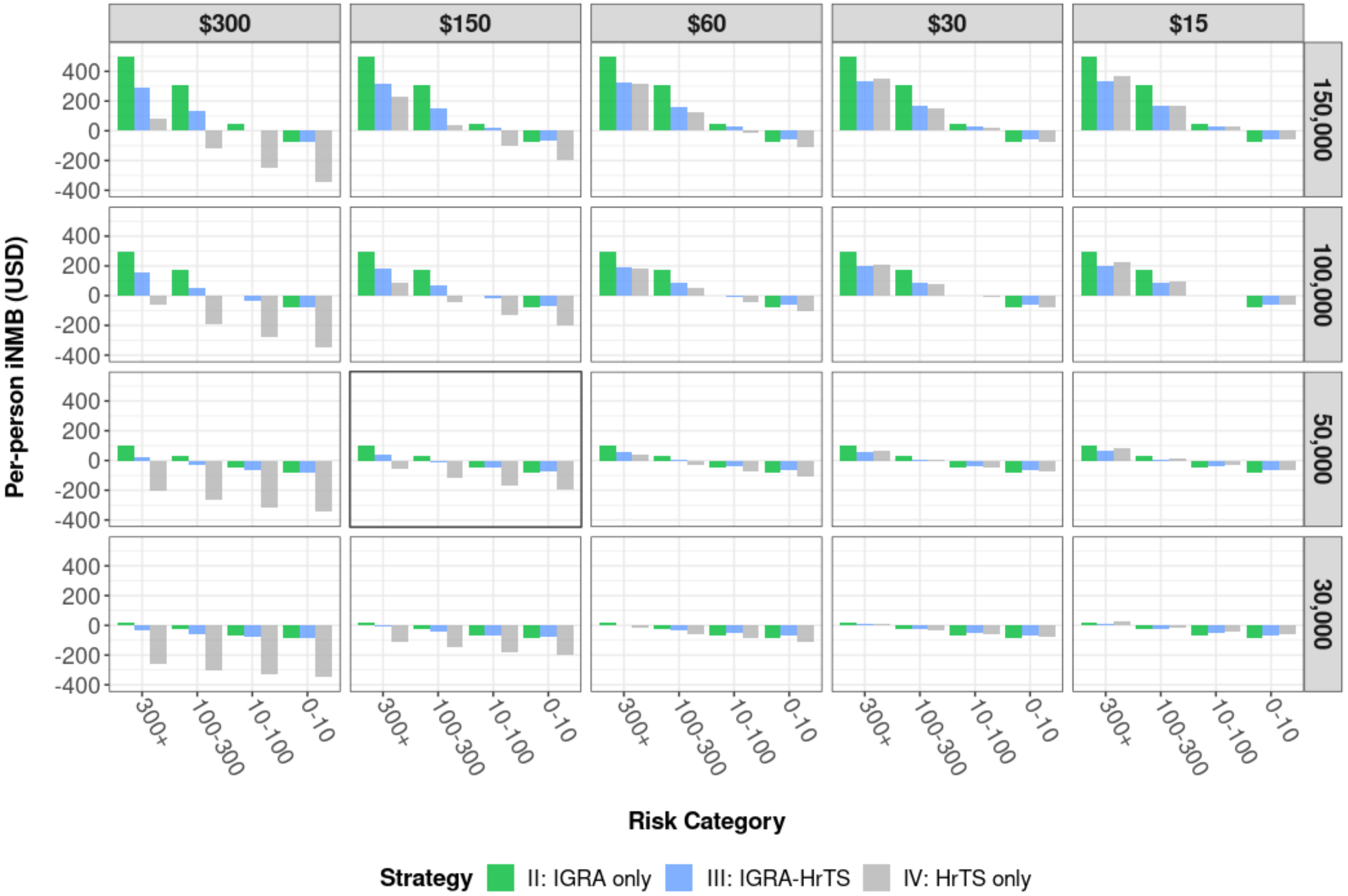

**Figure E2-1. Two-way sensitivity analysis of cost of the signature and willingness-to-pay threshold on the per-person incremental net monetary benefit relative to Strategy I (no screening), in the healthcare sector perspective.** The risk categories are epidemiological categorization of migrant populations based on TB incidence per 100k in 2019 for their country-of-origin.

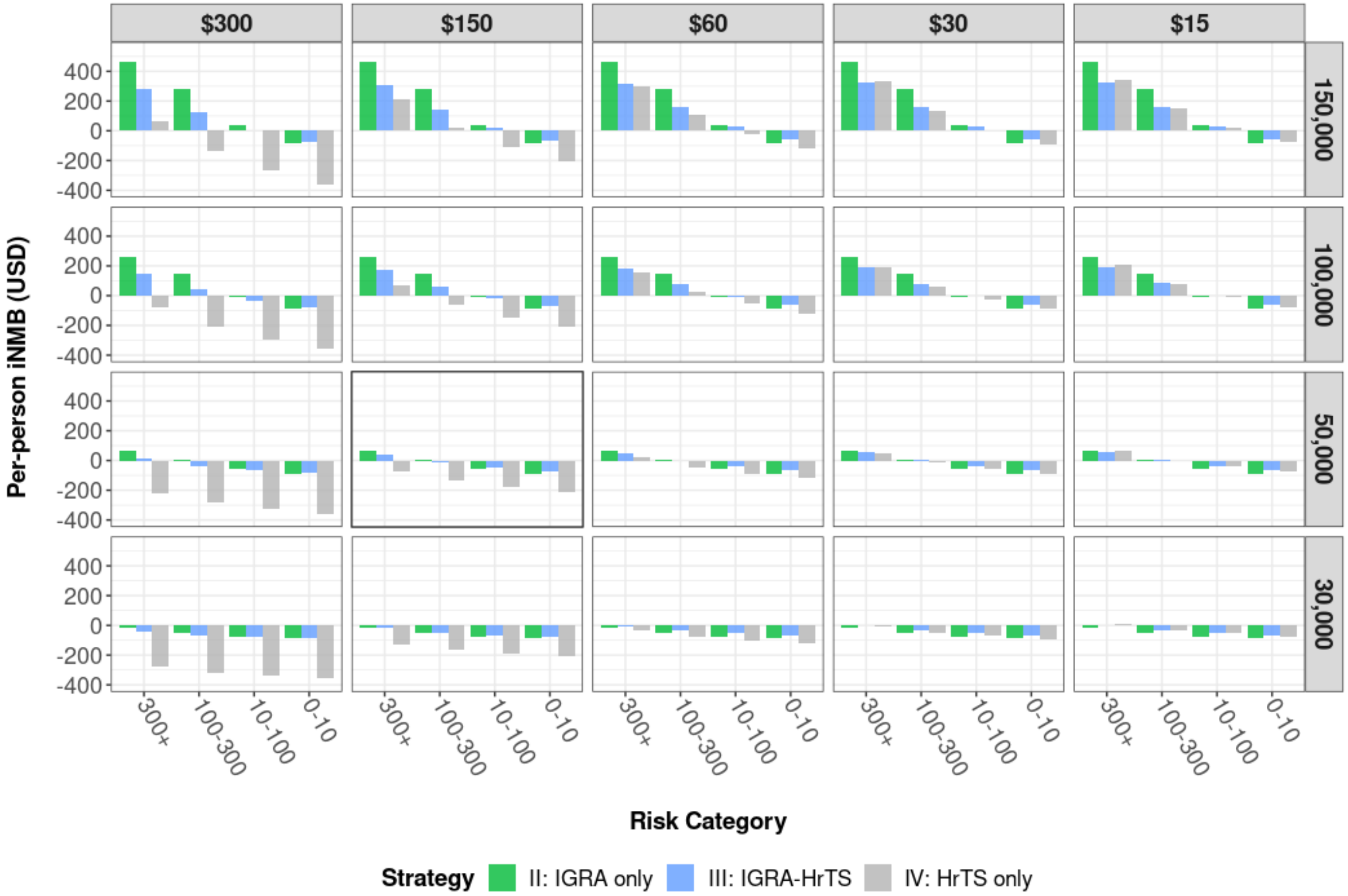

**Figure E2-2. Two-way sensitivity analysis of cost of the signature and willingness-to-pay threshold on the per-person incremental net monetary benefit relative to Strategy I (no screening), in the societal perspective.** The risk categories are epidemiological categorization of migrant populations based on TB incidence per 100k in 2019 for their country-of-origin.

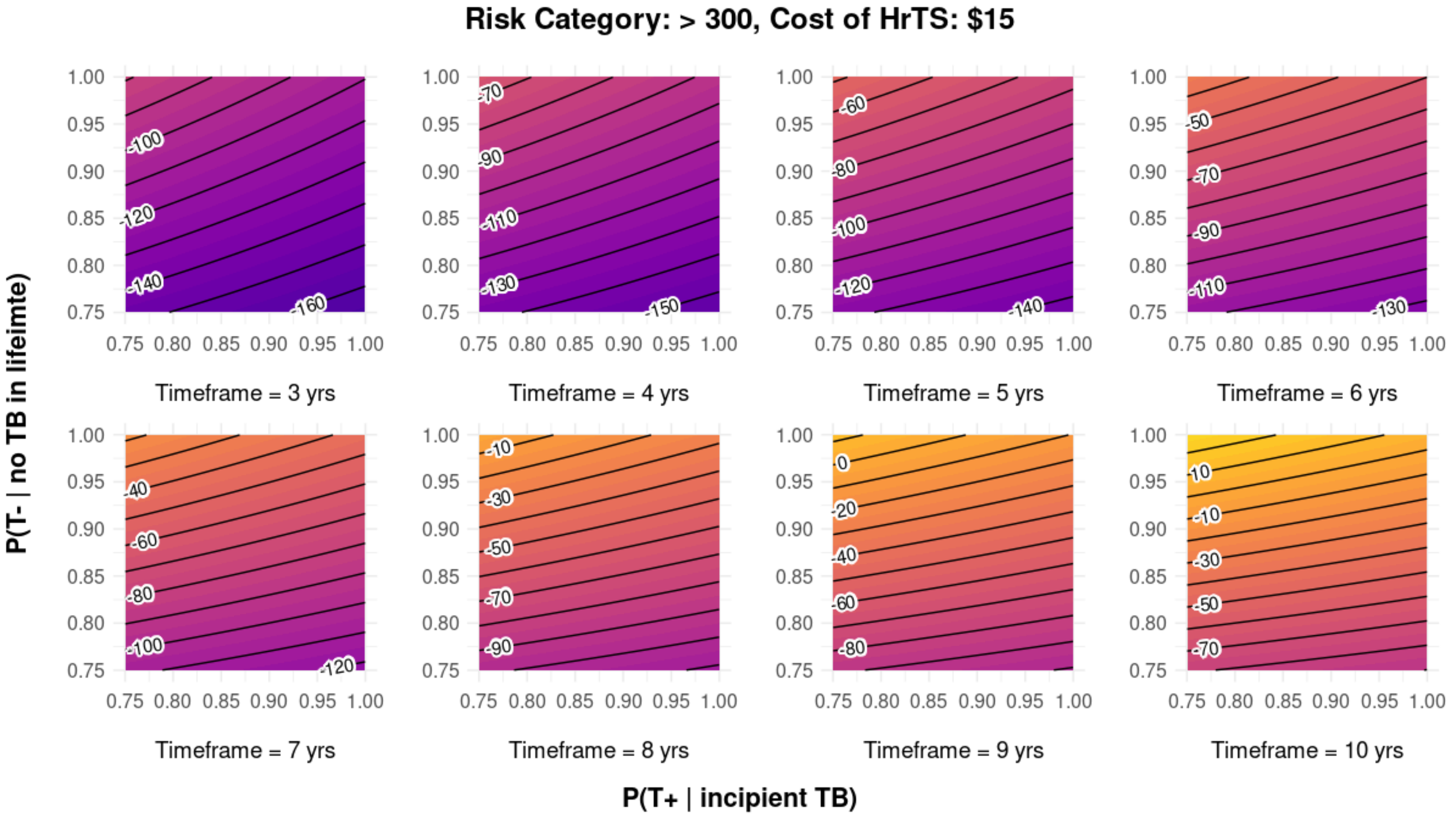

**Figure E3-1-1. Four-way sensitivity of the cost, sensitivity, and specificity of the RNA test, with the economic evaluation conducted in the healthcare sector perspective.** The contours represent the incremental net monetary benefit of Strategy IV (HrTS-only) to Strategy II (IGRA-only), for the highest risk group (country-of-origin TB incidence > 300 cases / 100k population in 2019) at the lowest cost of HrTS. Negative values indicate that Strategy IV is dominated by Strategy II. Willingness-to-pay threshold is 150,000 USD/ QALY gained.

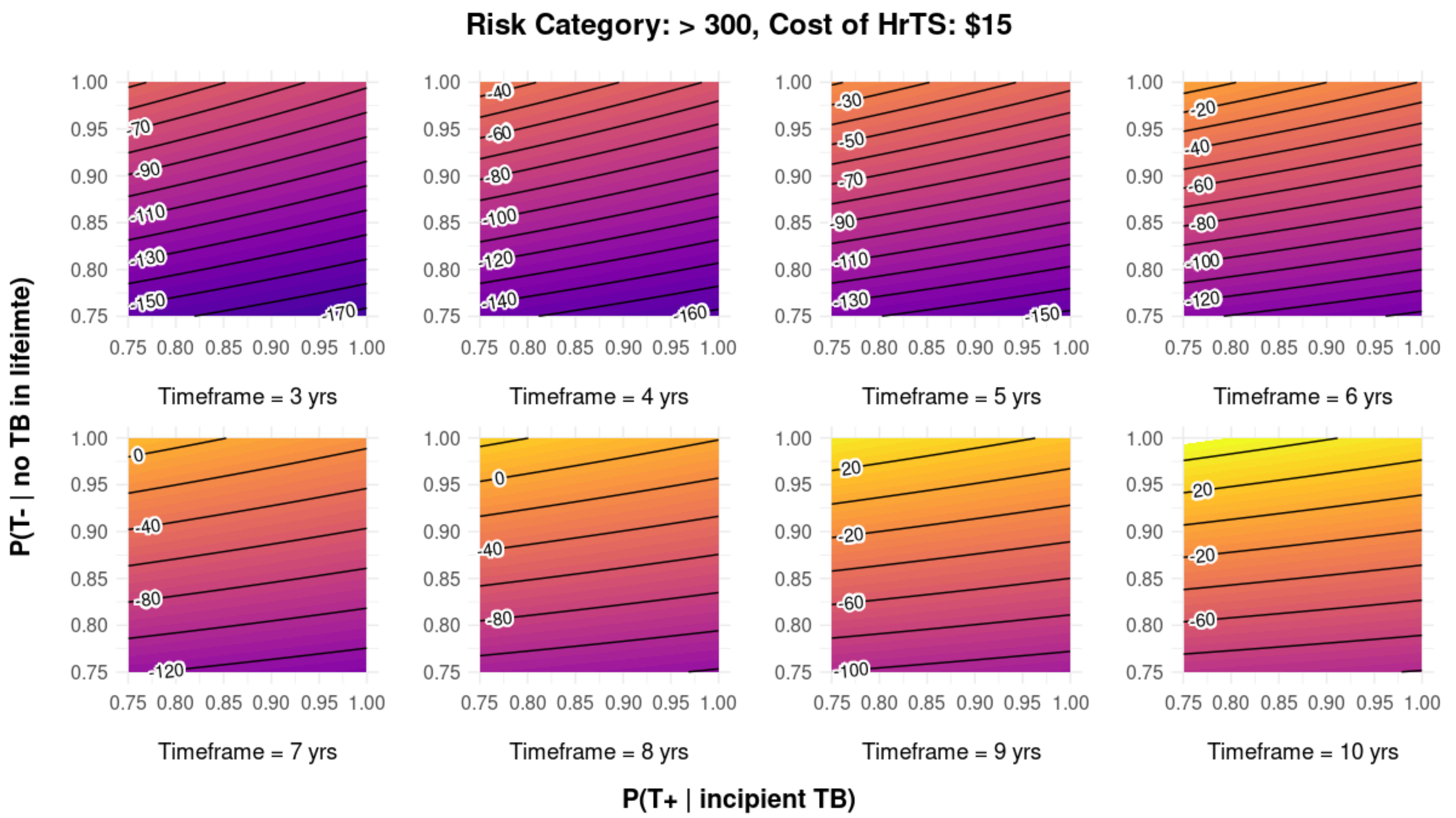

**Figure E3-1-2. Four-way sensitivity of the cost, sensitivity, and specificity of the HrTS, with the economic evaluation conducted in the societal perspective.** The contours represent the incremental net monetary benefit of Strategy IV (HrTS-only) to Strategy II (IGRA-only), for the highest risk group (country-of-origin TB incidence > 300 cases / 100k population in 2019) at the lowest cost of HrTS. Negative values indicate that Strategy IV is dominated by Strategy II. Willingness-to-pay threshold is 150,000 USD/ QALY gained.

#### Appendix 10

##### Alternative scenario for more rapid decline in TB progression risk

Description of scenario: As post-arrival screening program can only avert cases from *Mtb* infection acquired prior to the initial U.S. entry, we conducted a scenario analysis where we relaxed the assumption that all future TB cases were due to reactivation from prior *Mtb* infection.

In this scenario, we assumed a more rapid decline in the rate of progression to TB disease among those who arrived in the U.S. with *Mtb* infection. In this scenario, the annual number of TB cases for each future year was assumed to remained the same as in the main analysis, but the proportion of cases attributable to a pre-existing TB infection decreased over time. The remainder of the cases were assumed to be due to (re)infection after U.S. entry. We assumed the TB reactivation risk declined at a rate such that only 70% of the TB cases in year 10 were from reactivation, and 50% in year 20. This was operationalized as a 3.4% annual decline in the fraction of cases in a given year that were attributable to pre-existing infection.

#### Table E10. Testing and treatment outcomes for alternative scenario assuming more rapid decline in rates of progression to TB disease following U.S. entry

| | PPV for future TB disease (%, 95% CI) | | NPV for future TB disease (%, 95% CI) | | Percentage of population treated, by regimen type (%, 95% CI) | | | Reduction in TB cases (%, 95% CI) |
|---|---|---|---|---|---|---|---|---|
| | Within 2 years | Over lifetime | Within 2 years | Over lifetime | *Mtb* infection | Incipient TB | TB disease | |
| **Whole cohort** | | | | | | | | |
| ***No screening*** | NA | NA | NA | NA | NA | NA | 0.27 (0.24, 0.30) | *ref* |
| ***IGRA only*** | 0.4 (0.3, 0.5) | 1.2 (1.0, 1.4) | 100 (100, 100) | 100 (99.9, 100) | 8.5 (7.0, 10.3) | NA | 0.21 (0.18, 0.25) | 20.6 (17.0, 25.5) |
| ***IGRA-HrTS*** | 3.5 (2.9, 4.0) | 4.4 (3.6, 5.1) | 100 (100, 100) | 99.9 (99.9, 99.9) | NA | 1.0 (0.8, 1.2) | 0.25 (0.21, 0.28) | 8.1 (7.4, 9.2) |
| ***HrTS only*** | 0.5 (0.4, 0.5) | 0.6 (0.6, 0.7) | 100 (100, 100) | 99.9 (99.9, 99.9) | NA | 8.8 (7.8, 9.6) | 0.24 (0.21, 0.28) | 10.6 (9.4, 12.2) |
| **Risk category I** | | | | | | | | |
| ***No screening*** | NA | NA | NA | NA | NA | NA | 0.91 (0.81, 1.01) | *ref* |
| ***IGRA only*** | 0.9 (0.8, 1.1) | 3.1 (2.6, 3.5) | 99.9 (99.9, 100) | 99.8 (99.8, 99.9) | 11.3 (9.6, 13.1) | NA | 0.72 (0.60, 0.84) | 20.2 (16.9, 24.8) |
| ***IGRA-HrTS*** | 8.2 (6.9, 9.7) | 10.5 (9.0, 12.1) | 99.9 (99.9, 100) | 99.6 (99.5, 99.6) | NA | 1.4 (1.1, 1.6) | 0.83 (0.73, 0.93) | 8.7 (8.1, 10.0) |
| ***HrTS only*** | 1.6 (1.4, 1.9) | 2.1 (1.9, 2.4) | 100 (100, 100) | 99.6 (99.6, 99.6) | NA | 9.0 (8.0, 9.8) | 0.80 (0.70, 0.90) | 11.6 (9.9, 12.8) |
| **Risk category II** | | | | | | | | |
| ***No screening*** | NA | NA | NA | NA | NA | NA | 0.51 (0.44, 0.58) | *ref* |
| ***IGRA only*** | 0.7 (0.6, 0.8) | 2.0 (1.7, 2.3) | 100 (100, 100) | 99.9 (99.9, 100) | 10.3 (8.7, 12.1) | NA | 0.40 (0.32, 0.48) | 22.8 (18.4, 28.8) |
| ***IGRA-HrTS*** | 5.8 (4.7, 6.7) | 7.5 (6.0, 8.7) | 100 (100, 100) | 99.8 (99.7, 99.8) | NA | 1.3 (1.0, 1.5) | 0.47 (0.41, 0.53) | 9.2 (8.4, 9.5) |
| ***HrTS only*** | 1.0 (0.8, 1.1) | 1.2 (1.0, 1.4) | 100 (100, 100) | 99.8 (99.7, 99.8) | NA | 8.9 (7.9, 9.7) | 0.46 (0.40, 0.53) | 10.7 (9.5, 11.2) |
| **Risk category III** | | | | | | | | |
| ***No screening*** | NA | NA | NA | NA | NA | NA | 0.20 (0.17, 0.22) | *ref* |
| ***IGRA only*** | 0.3 (0.2, 0.3) | 0.9 (0.7, 1.0) | 100 (100, 100) | 100 (100, 100) | 8.4 (6.9, 10.1) | NA | 0.16 (0.13, 0.19) | 19.4 (16.2, 23.7) |
| ***IGRA-HrTS*** | 2.5 (2.1, 2.9) | 3.3 (2.6, 3.7) | 100 (100, 100) | 99.9 (99.9, 99.9) | NA | 1.0 (0.8, 1.2) | 0.18 (0.16, 0.21) | 7.3 (6.5, 8.7) |
| ***HrTS only*** | 0.4 (0.3, 0.4) | 0.4 (0.4, 0.5) | 100 (100, 100) | 99.9 (99.9, 99.9) | NA | 8.8 (7.8, 9.5) | 0.18 (0.15, 0.20) | 10.3 (9.3, 12.6) |
| **Risk category IV** | | | | | | | | |
| ***No screening*** | NA | NA | NA | NA | NA | NA | 0.01 (0.01, 0.02) | *ref* |
| ***IGRA only*** | 0.03 (0.02, 0.04) | 0.1 (0.1, 0.2) | 100 (100, 100) | 100 (100, 100) | 4.8 (3.7, 6.5) | NA | 0.01 (0.01, 0.02) | 17.4 (7.4, 33.3) |
| ***IGRA-HrTS*** | 0.3 (0.2, 0.4) | 0.3 (0.2, 0.4) | 100 (100, 100) | 99.9 (99.9, 99.9) | NA | 0.6 (0.4, 0.8) | 0.01 (0.01, 0.02) | 6.1 (2.5, 14.3) |
| ***HrTS only*** | 0.03 (0.03, 0.04) | 0.03 (0.03, 0.04) | 100 (100, 100) | 99.9 (99.9, 100) | NA | 8.6 (7.6, 9.4) | 0.01 (0.01, 0.02) | 7.7 (2.5, 14.3) |

As defined in the main text, risk categories are the epidemiological categorization of migrant populations based on TB incidence per 100k in 2019 for their country-of-origin*:* risk category I (≥300); risk category II (100-300), risk category III (10-100), risk category IV (0-10).

**Table E11. Cost-effectiveness results for alternative scenario assuming more rapid decline in rates of progression to TB disease following U.S. entry**

| Strategy | TB related HC costs | Other HC expenditures | TB related non-HC costs | Other non-HC expenditures | Productivity gain[1] | Total Costs[2] $ | Total QALY gain | Inc. Cost[2] | Inc. Effectiveness[3] (QALYs) | ICER | Inc. NMB[4] |
|---|---|---|---|---|---|---|---|---|---|---|---|
| **Healthcare Sector Perspective** | | | | | | | | | | | |
| **IGRA-HrTS** | 71.6 (55.9, 89.6) | 0.8 (0.4, 1.3) | -- | -- | -- | 72.5 (56.6, 90.5) | 0.00073 (0.00042, 0.00107) | 72.5 | 0.00073 | 99,315 | 37.0 |
| **IGRA only** | 95.9 (72.5, 125.8) | 2.6 (1.3, 3.8) | -- | -- | -- | 98.5 (74.0, 127.8) | 0.00098 (0.00066, 0.00133) | 26.0 | 0.00025 | 104,000 | 48.5 |
| **HrTS only** | 76.1 (56.1, 102.3) | 1.0 (0.5, 1.5) | -- | -- | -- | 77.1 (57.1, 103.3) | 0.00068 (0.00034, 0.00105) | *NA* | *NA* | Dominated | 24.9 |
| **Societal Perspective** | | | | | | | | | | | |
| **IGRA-HrTS** | 71.6 (55.9, 89.6) | 0.8 (0.4, 1.3) | 1.2 (0.6, 1.8) | 4.0 (2.0, 6.0) | 4.2 (1.2, 7.6) | 73.4 (57.5, 91.6) | 0.00073 (0.00042, 0.00107) | 73.4 | 0.00073 | 100,548 | 36.1 |
| **IGRA only** | 95.9 (71.5, 125.8) | 2.6 (1.3, 3.8) | 7.6 (5.3, 10.2) | 10.0 (4.9, 14.5) | 7.0 (2.9, 9.8) | 109.2 (84.4, 139.5) | 0.00098 (0.00066, 0.00133) | 35.8 | 0.00025 | 143,200 | 37.8 |
| **HrTS only** | 76.1 (56.1, 102.3) | 1.0 (0.5, 1.5) | 12.6 (10.1, 15.4) | 4.5 (2.3, 6.9) | 4.7 (1.5, 8.3) | 89.6 (69.2, 115.7) | 0.00068 (0.00034, 0.00105) | *NA* | *NA* | Dominated | 12.4 |

Notes: HC, healthcare; QALY, quality-adjusted life years; ICER, incremental cost-effectiveness ratio; NMB, net monetary benefit

*1* All costs, expenditures, productivity gain, and QALY gain were estimated relative to the "No Screening" strategy.

*2* Productivity gain was attributable to mortality aversion.

*3* For analysis in the healthcare sector perspective, Total Costs = (TB related healthcare costs + Other healthcare expenditures); for analysis in the societal perspective, Total Costs = (TB related healthcare costs + Other healthcare expenditures + TB related non-healthcare costs + Other non-healthcare expenditures – Productivity gain)

*4* The Inc. cost and Inc. effectiveness in columns 9-10 were estimated relative to the next best strategy.

*5* The NMB were based on $150,000/QALY; Inc. NMB = (Inc. effectiveness* $150,000/QALY) – Inc. costs

#### Appendix 11

##### Data Dictionary

This data dictionary describes the variables in the regression of the fitted TB risk model reported in the study “Hill AN, Cohen T, Salomon JA, Menzies NA. High-resolution estimates of tuberculosis incidence among non-U.S.-born persons residing in the United States, 2000–2016. *Epidemics* 2020; **33**: 100419”, which was funded by the U.S. Centers for Disease Control and Prevention, National Center for HIV/AIDS, Viral Hepatitis, STD, and TB Prevention Epidemiologic and Economic Modeling Agreement (#5NU38PS004644). The regression output was made available to the authors of this current project as an `.rdata` object. The R object is available at << Dataverse link will be added once the manuscript is accepted and finalized >>.

| Variable Name | Data Type | Description* |
|---|---|---|
| yse10r, yse20r, yse30r, yse40r, yse50r, yse60r | indicator variable | 1 if the number of years since entry rounded to the nearest decade fall into the indicated 10-year interval; 0 otherwise. |
| yoe2010r, yoe 2000r, yoe1990r, yoe1980r, yoe1970r, yoe1960r, yoe1950r | indicator variable | 1 if the year-of-entry rounded to the nearest decade fall into the indicated 10-year interval; 0 otherwise. |
| entry_this_year | indicator variable | 1 if the estimate is for TB risk in the entry year; 0 otherwise. |
| age_over_90 | indicator variable | 1 if the individual is older than 90 years old; 0 otherwise. |
| yoe_pre_1950 | indicator variable | 1 if the individual entered before year 1950; 0 otherwise. |
| years_since | continuous variable | Number of years since entry. |
| entry_age | continuous variable | Age at entry, top coded at age 91. |
| entry_year0 | continuous variable | De-meaned value for the entry year = 32.56782 + (entry_year – 2016) |
| origin | categorical variable | Country-of-origin, in ISO 3166-1 alpha-3 codes (ISO3) |
| (offset) | continuous variable | The population size of the stratum. |

* Further details given in the original article “Hill AN, Cohen T, Salomon JA, Menzies NA. High-resolution estimates of tuberculosis incidence among non-U.S.-born persons residing in the United States, 2000–2016. *Epidemics* 2020; **33**: 100419”.

4 Living Longer: Historical and Projected Life Expectancy in the United States, 1960 to 2060. .

5 R Core Team. R: A language and environment for statistical computing. R Foundation for Statistical Computing, Vienna, Austria. 2021. https://www.R-project.org/.

6 Lee-Rodriguez C, Wada PY, Hung Y-Y, Skarbinski J. Association of Mortality and Years of Potential Life Lost With Active Tuberculosis in the United States. *JAMA Netw Open* 2020; **3**: e2014481.

7 Stout JE, Wu Y, Ho CS, *et al.* Evaluating latent tuberculosis infection diagnostics using latent class analysis. *Thorax* 2018; **73**: 1062–70.

8 World Health Organization. Development of a Target Product Profile (TPP) and a framework for evaluation for a test for predicting progression from tuberculosis infection to active disease. 2017.

9 Treatment for Latent TB Infection | 2020 State and City TB Report | Data & Statistics | TB | CDC. 2022; published online Jan 18. https://www.cdc.gov/tb/statistics/indicators/2020/LatentTBInfection.htm (accessed June 21, 2022).

10 Sandul AL, Nwana N, Holcombe JM, *et al.* High Rate of Treatment Completion in Program Settings With 12-Dose Weekly Isoniazid and Rifapentine for Latent Mycobacterium tuberculosis Infection. *Clin Infect Dis* 2017; **65**: 1085–93.

11 Sterling TR, Moro RN, Borisov AS, *et al.* Flu-like and Other Systemic Drug Reactions Among Persons Receiving Weekly Rifapentine Plus Isoniazid or Daily Isoniazid for Treatment of Latent Tuberculosis Infection in the PREVENT Tuberculosis Study. *Clin Infect Dis Off Publ Infect Dis Soc Am* 2015; **61**: 527–35.

12 Zenner D, Beer N, Harris RJ, Lipman MC, Stagg HR, van der Werf MJ. Treatment of Latent Tuberculosis Infection. *Ann Intern Med* 2017; **167**: 248–55.

13 Hamada Y, Paulos L, Baruch NG, Cronin W. Proposed Approach for 4-Month Treatment of Culture-Negative Pulmonary Tuberculosis in Adults. *Ann Am Thorac Soc* 2016; **13**: 1657–8.

14 Guo N, Marra F, Marra CA. Measuring health-related quality of life in tuberculosis: a systematic review. *Health Qual Life Outcomes* 2009; **7**: 14.

15 Jo Y, Shrestha S, Gomes I, *et al.* Model-based Cost-effectiveness of State-level Latent Tuberculosis Interventions in California, Florida, New York, and Texas. *Clin Infect Dis Off Publ Infect Dis Soc Am* 2021; **73**: e3476–82.

16 Menzies NA, Quaife M, Allwood BW, *et al.* Lifetime burden of disease due to incident tuberculosis: a global reappraisal including post-tuberculosis sequelae. *Lancet Glob Health* 2021; **9**: e1679–87.

17 CDC Estimates for LTBI Treatment Costs | Publications & Products | TB | CDC. 2022; published online April 13. https://www.cdc.gov/tb/publications/infographic/ltbi-treatment-costs.htm (accessed June 22, 2022).

18 Infographics | Publications & Products | TB | CDC. 2021; published online Oct 25. https://www.cdc.gov/tb/publications/infographic/appendix.htm (accessed June 22, 2022).

19 Clinical Laboratory Fee Schedule | CMS. https://www.cms.gov/medicare/medicare-fee-for-service-payment/clinicallabfeesched (accessed Feb 19, 2023).

20 Article - Billing and Coding: Chest X-Ray Policy (A57497). https://www.cms.gov/medicare-coverage-database/view/article.aspx?articleId=57497 (accessed Feb 19, 2023).

21 Jiao B, Basu A. Catalog of Age- and Medical Condition—Specific Healthcare Costs in the United States to Inform Future Costs Calculations in Cost-Effectiveness Analysis. *Value Health* 2021; **24**: 957–65.

22 U.S. Bureau of Labor Statistics. Age of reference person: Annual expenditure means, shares, standard errors, and coefficients of variation, Consumer Expenditure Surveys, 2021. https://www.bls.gov/cex/tables/calendar-year/mean-item-share-average-standard-error/reference-person-age-ranges-2021.pdf (accessed April 10, 2023).

23 Grosse SD, Krueger KV, Pike J. Estimated annual and lifetime labor productivity in the United States, 2016: implications for economic evaluations. *J Med Econ* 2019; **22**: 501–8.

24 Neumann PJ, Ganiats TG, Russell LB, Sanders GD, Siegel JE, editors. Cost-Effectiveness in Health and Medicine. Oxford University Press, 2016 DOI:10.1093/acprof:oso/9780190492939.001.0001.
